# Lipidomics Identifies HFpEF Phenogroups and a High-Risk Metabolic Signature - The BElgian and CAnadian MEtabolomics in HFpEF (BECAME-HF) project

**DOI:** 10.64898/2026.03.31.26349865

**Authors:** Julie Hussin, Nassiba Menghoum, Anik Forest, Pamela Mehanna, Olivier Tastet, Julie Thompson Legault, Isabelle Frayne Robillard, Sibille Lejeune, David Vancraeynest, Clotilde Roy, Galadriel Briere, Gabrielle Boucher, Luc Bertrand, Sandrine Horman, David Rhainds, Jean-Claude Tardif, Christophe Beauloye, Anne-Catherine Pouleur, Christine Des Rosiers

**Author notes:** Contributed equally and are joint first authors (Julie Hussin, Nassiba Menghoum).

## Abstract

**Rationale:** Heart failure with preserved ejection fraction (HFpEF) is a heterogeneous syndrome with substantial unmet diagnostic and therapeutic needs. Circulating lipid metabolism is increasingly implicated in HFpEF pathophysiology but has not been systematically leveraged for molecular stratification.

**Objective:** To determine whether plasma lipidomics can identify molecular phenogroups of HFpEF associated with distinct clinical characteristics and outcomes.

**Methods and Results:** Untargeted plasma lipidomics was performed in non-HF subjects and HFpEF patients from a primary Belgian cohort and an independent Canadian cohort (n=177 in each cohort). In the Belgian cohort, 235 unique lipids spanning 19 subclasses were annotated, including 96 significantly associated with HFpEF (*q*<0.02). Unsupervised analyses revealed marked lipidomic heterogeneity, with a distinct HFpEF subgroup separable from non-HF subjects. Hierarchical clustering identified three phenogroups with divergent lipid profiles and clinical features. One phenogroup exhibited severe atrial dysfunction, congestion-related biomarkers, elevated indices of cardiac and liver fibrosis, and markedly reduced survival, a second was characterized by prominent metabolic syndrome features, and a third by preserved renal function. Cross-cohort comparison using a supervised classifier trained on 158 shared lipids confirmed analogous lower-risk phenogroups in the Canadian cohort, while the high-risk phenotype was underrepresented. A signature of 10 lipids across six subclasses, including long-chain acylcarnitines, ether phosphatidylcholines, and oxidized sphingomyelins, discriminated the high-risk group and correlated with markers of disease severity.

**Conclusion:** Our findings demonstrate that HFpEF comprises metabolically distinct patient subgroups across cohorts, revealing specific lipidomic dysfunctions that deepen our understanding of HFpEF heterogeneity and underlying pathophysiology.

## 1. Introduction

Heart failure with preserved ejection fraction (HFpEF) is a growing public health concern with high morbidity and mortality rates^1,2^. While heart failure (HF) affects 2-3% of the world’s population, more than half of all HF patients have HFpEF, with prevalence rising particularly in Western societies^3,4^, driven by an aging population and risk factors such as obesity, hypertension, and diabetes. Despite this increasing burden, management remains challenging, and SGLT2 inhibitors are the first therapeutic class proven to improve clinical outcomes in HFpEF, as demonstrated in pivotal trials like EMPEROR-Preserved and DELIVER^5,6^.

A major challenge in developing effective therapies is the marked phenotypic heterogeneity of HFpEF patients^7,8^. Patients present with diverse combinations of systemic inflammation that may lead to endothelial and microvascular dysfunction and, ultimately, cardiac structural abnormalities. Furthermore, HFpEF often presents with metabolic dysregulation, including hyperglycemia, hypertriglyceridemia, insulin resistance and metabolic-dysfunction-associated fatty liver disease (MAFLD)^9,10^. These metabolic disturbances are increasingly recognized as a major contributor to HFpEF pathophysiology, impairing myocardial energetic flexibility and promoting oxidative stress and mitochondrial dysfunction^11,12^. However, their mechanistic relevance across HFpEF subgroups remains poorly understood.

Over the past decade, phenomapping approaches using machine learning have emerged as powerful strategies to address this heterogeneity and better understand HFpEF pathophysiology. Pioneering work by Shah et al.^13^ using clinical and echocardiographic data identified three HFpEF phenogroups with distinct demographic and clinical characteristics, namely “metabolic obese”, “older vascular ageing” and “relatively younger low BNP” phenotypes, the former phenotype being the most characterized with respect to the metabolic involvement^14^. Subsequent studies have consistently demonstrated that HFpEF comprises biologically distinct subtypes with different outcomes^15–19^. More recent efforts incorporating proteomics within PROMIS-HFpEF^20^ have provided initial mechanistic insights, particularly regarding inflammation and fibrosis.

Given the strong metabolic signature of HFpEF, metabolomics represents a promising tool to further dissect its heterogeneity. However, previous studies have mostly relied on targeted analyses of a limited number of metabolites such as acylcarnitines and other energy-related metabolites^21–24^ and have not captured the complexity of metabolic alterations in HFpEF phenogroups. Recent small human studies using untargeted mass spectrometry profiling support the existence of circulating metabolic signatures in HFpEF but remain limited in scale and have not resolved patient heterogeneity into clinically actionable subgroups^25^. Furthermore, while some studies have identified lipid alterations in plasma and myocardial tissue from HF patients, these analyses have largely been limited to targeted approaches on a small number of lipids, primarily in patients with heart failure with reduced ejection fraction (HFrEF)^26–28^. As a result, the full breadth of lipid dysregulation in HFpEF remains unexplored, and no study to date has used untargeted lipidomics to define metabolically driven HFpEF phenogroups.

Given the central role of lipid dyshomeostasis in HFpEF pathophysiology^11,12^, we hypothesized that a discovery-based characterization of the plasma lipidome could reveal distinct HFpEF phenogroups and provide mechanistic insight into their underlying metabolic pathways. Clinical trial lipidomic data suggest that SGLT2 inhibition modulates multiple circulating lipid classes in HF patients, reinforcing lipids as both mechanistic readouts and potential stratifiers^29,30^. In the current study, we applied a comprehensive untargeted lipidomics workflow combined with unsupervised machine learning to identify clinically relevant HFpEF subgroups and evaluate whether specific lipid signatures correspond to unique phenotypes, pathophysiological mechanisms, and clinical outcomes. We identified phenogroups associated with distinct clinically relevant characteristics and outcomes, shedding light on their specific lipid-related metabolic dysregulations.

## 2. Methods

### Ethical statement

The study was conducted in accordance with the Declaration of Helsinki and approved by local ethics committees in Belgium (Comité d’éthique Hospitalo-Facultaire, Cliniques universitaires Saint-Luc and UCLouvain; approvals 2012/23AVR/199 and 2018/19MAR/118) and Canada (Montreal Heart Institute protocol #2017-2316). All participants provided written informed consent.

### Cohorts and clinical phenotyping

This study analyzed participants from two cohorts within the BECAME-HF program: a primary cohort (BECAME-HF1) and an independent secondary cohort (BECAME-HF2), referred to here as the Belgian and Canadian cohorts, respectively, according to their recruitment sites. This nomenclature is used solely for clarity and does not imply that geographic origin is a focus of the analysis. Detailed inclusion and exclusion criteria, as well as baseline demographic, clinical, and biochemical characteristics, have been previously reported^31^. The Belgian cohort is a single-center, real-world cohort of consecutive patients evaluated for HFpEF (Supplementary Text). The present study included 105 HFpEF patients and 72 age- and sex-matched non-HF subjects from that cohort. Non-HF subjects had no history of cardiovascular events but could present common cardiovascular risk factors. HFpEF patients were prospectively followed with structured ambulatory visits and/or telephone contacts every six months, allowing longitudinal assessment of mortality and heart-failure hospitalization. The Canadian cohort includes participants recruited from the Montreal Heart Institute (MHI) Biobank, comprising 74 HFpEF patients and 103 age- and sex-matched non-HF subjects. This cohort was used exclusively for cross-cohort comparisons.

In addition to standard clinical and laboratory measurements, we evaluated several established circulating biomarkers and composite scores capturing key pathophysiological domains relevant to HFpEF. These included markers of congestion (NT-proBNP, carbohydrate antigen 125 [CA-125]), cardiac injury and fibrosis (troponin T, fibroblast growth factor-23 [FGF-23], and soluble suppression of tumorigenicity-2 [sST2]), hepatic dysfunction and fibrosis (Fibrosis-4 [FIB-4] index, NAFLD score, albumin, and aminotransferases ratio [AST/ALT]), renal function (estimated glomerular filtration rate [eGFR]), platelet indices (platelet count and mean platelet volume), and cardiometabolic status (HDL-cholesterol, triglycerides, metabolic syndrome, and the triglyceride-glucose [TyG] index as a surrogate of insulin resistance). These biomarkers and scores are routinely used in clinical and translational research to reflect disease severity and multisystem involvement in HFpEF. Detailed definitions, formulas, and clinical thresholds are provided in the Supplementary Text.

### Sample collection and metabolomics workflow

Blood samples were obtained by venipuncture in the fasting state and kept on ice until centrifugation at 3500 rpm for 10 minutes. Thereafter, aliquots of plasma or serum were stored at −80 °C until further metabolite analyses. Stratified randomization of samples was achieved according to potential confounding factors, namely age, sex, disease status and treatment, to minimize batch effects. Targeted profiling of amino acids and organic acids was performed in subsets of both cohorts using established isotope-dilution gas chromatography-mass spectrometry (GC-MS) workflows, as previously described^32,33^ with modifications (Supplementary Text). Untargeted lipidomic profiling was conducted using a previously validated semi-quantitative liquid chromatography-mass spectrometry (LC-MS) platform optimized for plasma and serum. The Belgian cohort was analyzed in two sequential phases (pilot and main study), while the Canadian cohort was analyzed independently in a later batch. Quality control procedures included repeated analysis of pooled plasma samples, blank injections, and internal standards to monitor signal stability, mass accuracy, and retention time reproducibility^34,35^. An overview of the untargeted lipidomics workflow is shown in **Supplementary Figure S1**.

### Lipidomic data processing and differential analysis

Raw MS data were processed with a workflow using Mass Hunter Qualitative, as previously described^34^. In brief, lipid features were extracted and aligned, normalized to account for technical variability, and corrected for batch effects prior to downstream analysis. Detailed quality control procedures and processing parameters (missingness, imputation and variability criteria) are provided in Supplementary Text. Lipid species were annotated based on tandem MS and an in-house reference database. When multiple features corresponded to the same lipid, the dominant feature was retained for analysis. Analyses were performed separately within each cohort, and a common subset of annotated lipids was used for cross-cohort comparisons between the Belgian and Canadian cohorts (Cross-cohort dataset). For differential analysis, log₂-transformed lipid intensities were modeled using linear regression, with HFpEF status as the primary predictor and age and sex included as covariates. Effect sizes were estimated as log₂ fold-changes, and statistical significance was assessed using false discovery rate (FDR) correction for multiple testing with Storey method. Sensitivity analyses incorporating additional clinical covariates were performed in the primary cohort (Supplementary Text). Relationships among HFpEF-associated lipids were further explored using correlation-based network analysis (Pearson correlation threshold of *r* > 0.6) to identify lipid clusters, visualized using the Fruchterman-Reingold algorithm with the *igraph* package in R. Reproducibility across analytical phases and cohorts was assessed using concordance of effect size estimates (Supplementary Text).

### Phenogroup discovery

Principal Component Analysis (PCA) was performed on the 235 annotated lipids using the *prcomp* function in R to reduce dimensionality by projecting the correlated lipid variables onto orthogonal components. Comparison with non-HF subjects through unsupervised analyses are described in the Supplementary Text. Hierarchical clustering was then performed on the PC-transformed data to identify subgroups among HFpEF patients in the Belgian cohort based on the 235 annotated lipids. The optimal number of clusters (n=3) was determined using the Ward’s aggregation criterion as implemented in the *FactoMineR* package. To assess the robustness of this unsupervised clustering solution, we additionally applied a consensus clustering framework (clustOMICS^36^), which yielded highly concordant results (Supplementary Text). Given this consistency, the three-cluster hierarchical solution was retained for downstream analyses. Differences in lipid levels across the three HFpEF clusters were assessed using the non-parametric Kruskal-Wallis test, with a significance threshold (*p* < 7 × 10⁻⁴) to account for multiple testing, followed by Wilcoxon rank-sum post-hoc pairwise comparisons (*p* < 0.02). Heatmaps were generated to visualize cluster-specific lipidomic patterns.

### Clinical and outcomes analyses across phenogroups

Continuous variables were expressed as mean ± SD if normally distributed or as median (25^th^ and 75^th^ percentiles) if not normally distributed. Categorical variables were expressed as counts and percentages. Biochemical parameters were log10-transformed to reduce skewness. Clinical, biochemical and imaging parameters were compared between HFpEF Belgian clusters using one-way analysis of variance (ANOVA) with homogeneity of variance assessment and post-hoc Tuckey test or Kruskal-Wallis test for quantitative variable, or chi-square test, when appropriate. P-value < 0.05 was considered significant. Kaplan Meier curves were used to assess survival probability from all-causes with or without hospitalization. Event-free and overall survival was estimated using log rank test. Multivariable Cox regression analysis was performed for the secondary endpoint using commonly assessed clinical parameters with a *p* < 0.10 (**Table 1**). The selected covariates were chosen not only based on statistical criteria but also on their established prognostic relevance, as consistently reported in our previous publications^8,31^. In case of collinearity of covariables (*r* > 0.50), only the strongest of two covariates was proposed for inclusion into the multivariable model. For targeted metabolites, data were log2-transformed and compared across patient clusters using the non-parametric Kruskal-Wallis test. A *p* < 0.05 was considered statistically significant, and significant overall effects were followed by pairwise Wilcoxon rank-sum tests with Bonferroni correction.

**Table 1.**
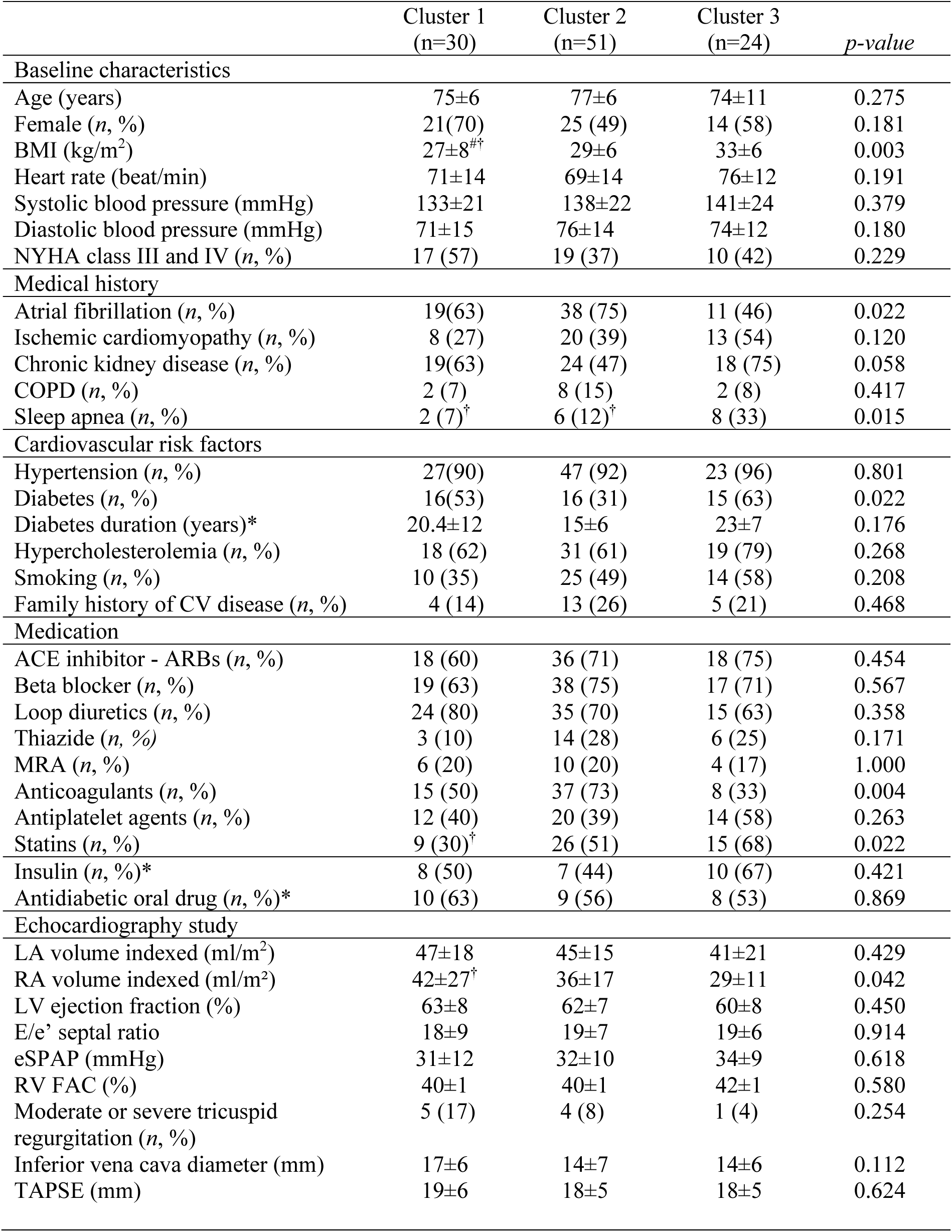
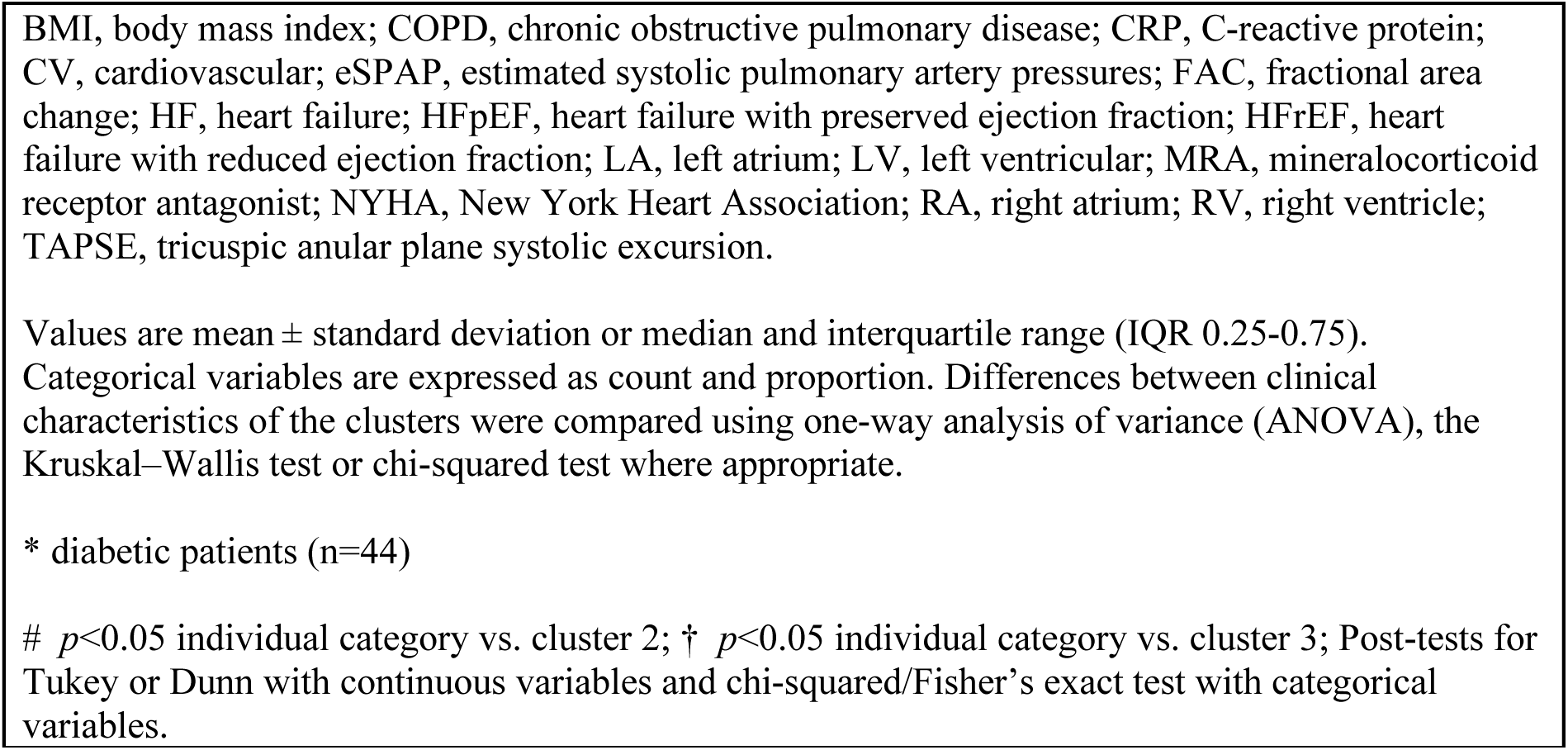
Baseline clinical characteristics of HFpEF patients according to cluster subgroups in the primary Belgian cohort.

|  | Cluster 1<br>(n=30) | Cluster 2<br>(n=51) | Cluster 3<br>(n=24) | *p-value* |
| --- | --- | --- | --- | --- |
| **Baseline characteristics** |  |  |  |  |
| Age (years) | 75±6 | 77±6 | 74±11 | 0.275 |
| Female (n, %) | 21(70) | 25 (49) | 14 (58) | 0.181 |
| BMI (kg/m<sup>2</sup>) | 27±8<sup>#†</sup> | 29±6 | 33±6 | 0.003 |
| Heart rate (beat/min) | 71±14 | 69±14 | 76±12 | 0.191 |
| Systolic blood pressure (mmHg) | 133±21 | 138±22 | 141±24 | 0.379 |
| Diastolic blood pressure (mmHg) | 71±15 | 76±14 | 74±12 | 0.180 |
| NYHA class III and IV (n, %) | 17 (57) | 19 (37) | 10 (42) | 0.229 |
| **Medical history** |  |  |  |  |
| Atrial fibrillation (n, %) | 19(63) | 38 (75) | 11 (46) | 0.022 |
| Ischemic cardiomyopathy (n, %) | 8 (27) | 20 (39) | 13 (54) | 0.120 |
| Chronic kidney disease (n, %) | 19(63) | 24 (47) | 18 (75) | 0.058 |
| COPD (n, %) | 2 (7) | 8 (15) | 2 (8) | 0.417 |
| Sleep apnea (n, %) | 2 (7)<sup>†</sup> | 6 (12)<sup>†</sup> | 8 (33) | 0.015 |
| **Cardiovascular risk factors** |  |  |  |  |
| Hypertension (n, %) | 27(90) | 47 (92) | 23 (96) | 0.801 |
| Diabetes (n, %) | 16(53) | 16 (31) | 15 (63) | 0.022 |
| Diabetes duration (years)* | 20.4±12 | 15±6 | 23±7 | 0.176 |
| Hypercholesterolemia (n, %) | 18 (62) | 31 (61) | 19 (79) | 0.268 |
| Smoking (n, %) | 10 (35) | 25 (49) | 14 (58) | 0.208 |
| Family history of CV disease (n, %) | 4 (14) | 13 (26) | 5 (21) | 0.468 |
| **Medication** |  |  |  |  |
| ACE inhibitor - ARBs (n, %) | 18 (60) | 36 (71) | 18 (75) | 0.454 |
| Beta blocker (n, %) | 19 (63) | 38 (75) | 17 (71) | 0.567 |
| Loop diuretics (n, %) | 24 (80) | 35 (70) | 15 (63) | 0.358 |
| Thiazide (n, %) | 3 (10) | 14 (28) | 6 (25) | 0.171 |
| MRA (n, %) | 6 (20) | 10 (20) | 4 (17) | 1.000 |
| Anticoagulants (n, %) | 15 (50) | 37 (73) | 8 (33) | 0.004 |
| Antiplatelet agents (n, %) | 12 (40) | 20 (39) | 14 (58) | 0.263 |
| Statins (n, %) | 9 (30)<sup>†</sup> | 26 (51) | 15 (68) | 0.022 |
| Insulin (n, %)* | 8 (50) | 7 (44) | 10 (67) | 0.421 |
| Antidiabetic oral drug (n, %)* | 10 (63) | 9 (56) | 8 (53) | 0.869 |
| **Echocardiography study** |  |  |  |  |
| LA volume indexed (ml/m<sup>2</sup>) | 47±18 | 45±15 | 41±21 | 0.429 |
| RA volume indexed (ml/m<sup>2</sup>) | 42±27<sup>†</sup> | 36±17 | 29±11 | 0.042 |
| LV ejection fraction (%) | 63±8 | 62±7 | 60±8 | 0.450 |
| E/e' septal ratio | 18±9 | 19±7 | 19±6 | 0.914 |
| eSPAP (mmHg) | 31±12 | 32±10 | 34±9 | 0.618 |
| RV FAC (%) | 40±1 | 40±1 | 42±1 | 0.580 |
| Moderate or severe tricuspid regurgitation (n, %) | 5 (17) | 4 (8) | 1 (4) | 0.254 |
| Inferior vena cava diameter (mm) | 17±6 | 14±7 | 14±6 | 0.112 |
| TAPSE (mm) | 19±6 | 18±5 | 18±5 | 0.624 |

BMI, body mass index; COPD, chronic obstructive pulmonary disease; CRP, C-reactive protein; CV, cardiovascular; eSPAP, estimated systolic pulmonary artery pressures; FAC, fractional area change; HF, heart failure; HFpEF, heart failure with preserved ejection fraction; HFrEF, heart failure with reduced ejection fraction; LA, left atrium; LV, left ventricular; MRA, mineralocorticoid receptor antagonist; NYHA, New York Heart Association; RA, right atrium; RV, right ventricle; TAPSE, tricuspid annular plane systolic excursion.
Values are mean ± standard deviation or median and interquartile range (IQR 0.25-0.75). Categorical variables are expressed as count and proportion. Differences between clinical characteristics of the clusters were compared using one-way analysis of variance (ANOVA), the Kruskal–Wallis test or chi-squared test where appropriate.
\* diabetic patients (n=44)

### Cross-cohort comparison

To evaluate the reproducibility of lipidomic phenogroups across cohorts, hierarchical clustering was applied to the Canadian cohort using the subset of 158 annotated lipids shared with the Belgian cohort (Cross-cohort dataset). To enable direct cross-cohort comparisons, lipidomic data from both cohorts were jointly visualized using PCA, after log2-transformation, batch effects correction and standardization. Hierarchical clustering was applied to the Canadian cohort on the Cross-cohort dataset. Similarity between phenogroups across cohorts was assessed by comparing cluster-averaged lipid profiles using correlation-based analyses. In addition, a supervised classification approach was used to test the transferability of cluster definitions across cohorts. A Random Forest classifier was trained on the Belgian cohort using a stratified 70/30 train-test split (Supplementary Text) and then applied to HFpEF patients from the Canadian cohort to predict cluster membership based on shared lipid features.

### Minimal lipid signature

To identify a minimal lipid signature associated with the high-risk HFpEF phenogroup in BECAME-HF1, we applied a resampling-based LASSO feature selection strategy, appropriate for high-dimensional and correlated lipidomic data (Supplementary Text). Lipids consistently selected across resampling iterations (>50%) were retained and combined in a multivariable ridge regression model to derive a continuous score reflecting similarity to the B1 cluster based on the selected lipid signature. Heatmap visualization of the retained lipids was performed using the *pheatmap* package in R with hierarchical clustering based on the Ward.D linkage method. We performed a network analysis based on pairwise correlations (r > 0.6) between annotated lipids measured in the BECAME-HF1 cohort and visualized it using the Fruchterman-Reingold algorithm with the *igraph* package in R. Associations between the resulting lipid signature and clinical variables (evaluated using correlation analyses adapted to variable type) were visualized using network_plot function in the *Corrr* package in R. Additional details on model fitting, resampling procedures, correlations and network-based visualization are provided in the Supplementary Text.

### Data and code availability

The lipidomic and metabolomic datasets generated and analyzed in this study are derived from human clinical cohorts. Processed lipidomic data matrices (log₂-transformed, normalized, and batch-corrected), including lipid annotations and cluster assignments, are available in a public repository. Summary-level data supporting the main findings of this study are provided in the Supplementary Tables. All custom code used for lipidomic data processing, statistical analyses, clustering, and visualization was written in R and is available on GitHub [https://github.com/HussinLab/BECAME_HF/] with appropriate documentation. Any additional information required to reanalyze the raw data is available from the corresponding authors upon reasonable request, subject to institutional data-sharing agreements and ethical approval.

## 4. Results

### Untargeted lipidomics reveals structured unique plasma lipid alterations in HFpEF

Baseline clinical and biochemical characteristics of non-HF subjects (n=72) and HFpEF patients (n=105) in the primary Belgian cohort (**Supplementary Table S1**) are consistent with those previously described for this cohort^31^. Initial targeted profiling of 21 amino acids and 12 organic acids revealed only marginal differences between HFpEF patients and non-HF subjects, consistent with prior studies^21^(**Supplementary Table S2**, Supplementary Text). We therefore applied untargeted lipidomics to identify plasma lipid features associated with HFpEF using our previously validated workflow^34,35^. After processing and quality control (workflow overview in **Supplementary Figure S1**), 1,666 lipid features were retained for downstream analyses. Adjusting for age and sex, 227 lipid features out of 1,666 were significantly associated with HFpEF (**Figure 1A-C**). Notably, these results were remarkably similar to those observed in our pilot study conducted on samples from the same cohort two years earlier (**Supplementary Figure S2**, Supplementary Text) supporting the robustness of our untargeted lipidomics workflow analysis^34^.

**Figure 1.**
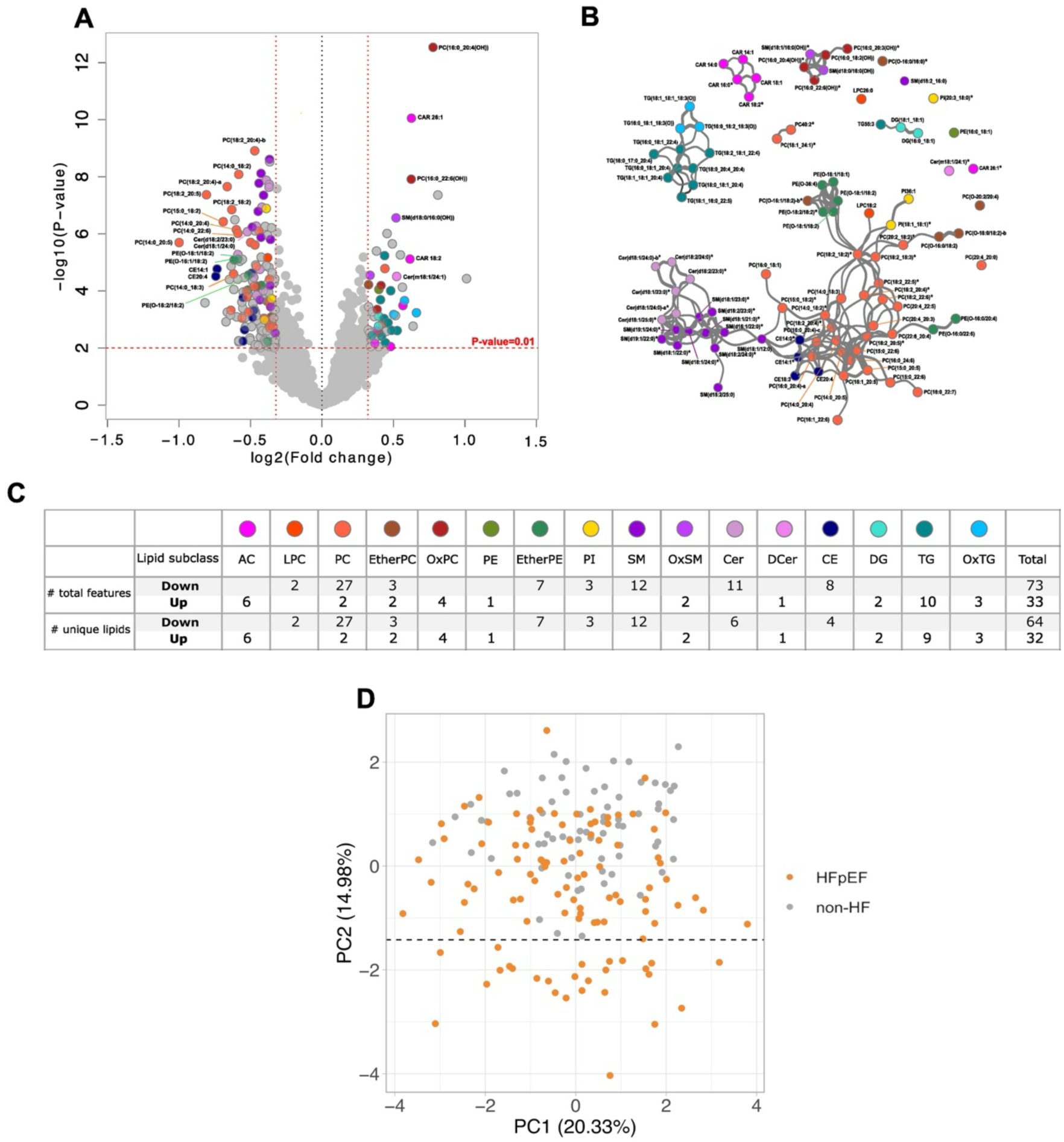
Untargeted plasma lipidomics reveals structured lipid alterations and heterogeneity in HFpEF. **(A)** Volcano plot showing plasma lipid features associated with HFpEF compared with non-HF subjects in the Belgian cohort after adjustment for age and sex. Annotated features with *p* < 0.01 (red line), |fold change| > 1.25 and Q < 0.02 are highlighted with different colors according to subclasses as described below in (C). **(B)** Lipid correlation network of HFpEF-associated annotated lipids (n = 96), constructed from positive Pearson correlations (*r* > 0.6) and visualized using a Fruchterman–Reingold layout, revealing four major correlation groups (I–IV) organized by lipid class or shared structural features. (*Lipid: lipids associated with HFpEF after adjustment for BMI, diabetes, eGFR, and NT-proBNP). **(C)** Distribution of the 96 annotated lipids significantly associated with HFpEF across 16 lipid subclasses (64 increased, 32 decreased). **(D)** Principal component analysis (PCA) based on the full set of 235 annotated lipids in non-HF subjects and HFpEF patients. A distinct subset of HFpEF patients (n = 27) separates along PC2 without overlap with non-HF subjects, using the most extreme PC2 value observed among non-HF subjects as the lower boundary.

Lipid annotation performed on the full dataset (Methods) yielded 235 unique annotated lipids spanning 19 subclasses, including 96 lipids out of 235, from 16 subclasses, significantly associated with HFpEF (64 increased and 32 decreased; **Figure 1C**; **Supplementary Excel File**) after adjustment for age and sex. A total of 43 remained associated after additional adjustment for BMI, diabetes, eGFR, and NT-proBNP (**Figure 1B**, **Supplementary Excel File**), with representation from acylcarnitines (CAR), sphingomyelins (SM), ether glycerophosphatidylcholine (PC) and glycerophosphatidylethanolamine (PE). As expected, correlation analysis on these 96 HFpEF-associated lipids revealed a structured organization into four major lipid correlation groups (**Figure 1B**, groups I–IV; ≥5 unique lipids) consisting of the same lipid (sub)class (Group I: long-chain acylcarnitnines (LC-CAR) but also spanning multiple subclasses (Group IV; ceramides (Cer) and SM with very-long-chain fatty acyl (VLC-FA)-containing side chains, PC and PE, ether PC and PE, and cholesterol esters (CE)) suggesting heterogeneity of lipid alterations among HFpEF patients. This was further investigated by principal component analysis (PCA) based on the 235 annotated lipids, which highlighted a distinct subset of 27 HFpEF patients separated along PC2 with no overlap with non-HF subjects (**Figure 1D**; **Supplementary Figure S3**; Supplementary Text). This indicate that a subset of HFpEF patients exhibits a globally distinct lipidomic profile relative to non-HF subjects. This separation is not explained by differences in BMI, with BMI adjustment having little impact on the overall HFpEF vs non-HF fold-changes (**Supplementary Figure S4A**). Furthermore, interaction analyses (**Supplementary Figure S4B**) suggested that BMI influences specific lipid features without driving the emergence of HFpEF subgroups. Altogether, these results reveal a distinct molecular subset within the HFpEF cohort that motivates downstream phenogrouping analyses.

### Plasma lipidomics delineates three metabolically distinct HFpEF phenogroups

To determine whether the lipidomic heterogeneity observed among HFpEF patients reflects discrete subphenotypes, we conducted hierarchical clustering on the 235 annotated plasma lipids in the Belgian cohort (**Figure 2A; Supplementary Table S2**). This analysis yielded a three-cluster solution, supported by consensus clustering (Supplementary Text). The resulting clusters displayed distinct global lipidomic patterns of changes across the various lipid subclasses that differed markedly from the aggregate HFpEF profile, consistent with phenotypic heterogeneity (**Figure 2B**; **Supplementary Excel File**). Notably, this Belgian cluster 1 (cluster B1) encompassed two-thirds of the HFpEF subset previously identified by PCA as lipidomically distinct (18 of 27 patients), whereas clusters B2 and B3 captured additional heterogeneity among the remaining HFpEF patients.

**Figure 2.**
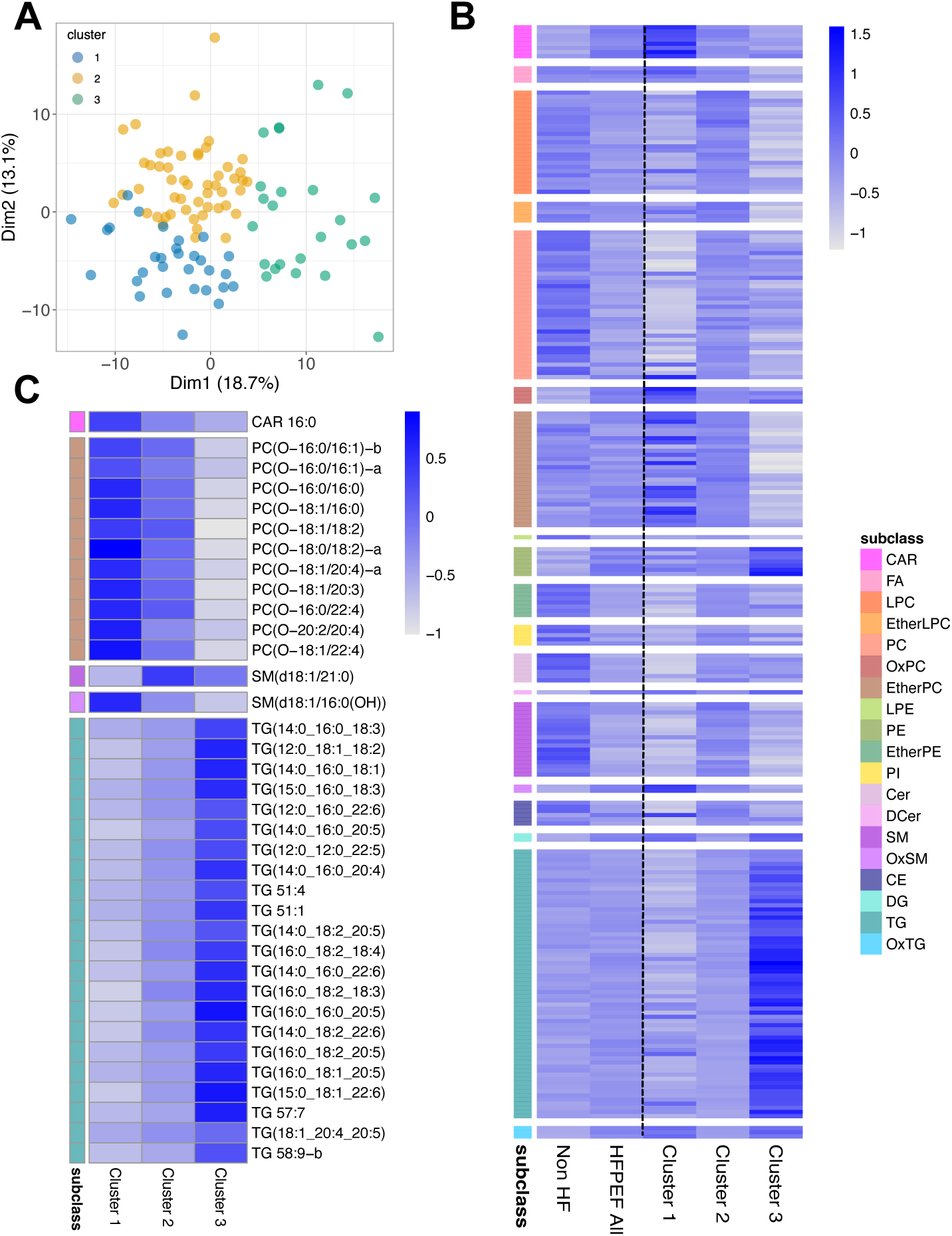
Plasma lipidomics identifies three metabolically distinct HFpEF phenogroups. **(A)** Hierarchical clustering of HFpEF patients based on 235 annotated plasma lipids in the Belgian cohort. Cluster assignment (B1–B3) is visualized on a PCA score plot, showing separation of patients according to lipidomic profiles. **(B)** Heatmap of the 235 annotated lipids across individual HFpEF clusters compared with the overall HFpEF population and non-HF subjects, illustrating distinct global lipidomic patterns for each phenogroup. **(C)** Heatmap of lipids significantly associated with individual HFpEF clusters, highlighting cluster-specific lipid signatures and opposing patterns across major lipid classes. Lipids are organized by subclass, and color intensity reflects relative abundance.

From the 235 annotated lipids, 158 (67%) differed significantly between clusters (Kruskal-Wallis *p* < 7 × 10^−4^ and Wilcoxon rank-sum post-hoc pairwise *p* <0.02; **Methods**), revealing cluster-specific lipid signatures in which most lipids within a given (sub)class changed in a consistent direction (**Supplementary Excel File**). Of these, 36 lipids showed significant differences across all pairwise cluster comparisons, while additional subsets of lipids distinguished individual clusters from the others (**Figure 2C, Supplementary Figure S5**). Notably, clusters B1 and B3 exhibited opposing lipidomic profiles across multiple lipid classes, with LC-CAR, ether PC and oxidized SM (OxSM) highest in cluster B1, whereas ether PC levels were lowest and most triglycerides (TGs) highest in cluster B3. Cluster B2 displayed more intermediate patterns and fewer distinguishing features, except for SM with VLCFA, lysoPC and specific PC, which showed the highest levels among clusters (**Supplementary Figure S5**). Together, these three molecularly distinct phenogroups provide a foundation for subsequent clinical and prognostic characterization.

### Clinical and prognosis differences between HFpEF phenogroups

To assess the clinical relevance of HFpEF phenogroups defined by plasma lipidomics, we compared baseline clinical and biochemical parameters across clusters (**Table 1-2**). Several baseline features differed significantly among clusters (**Table 1**; *p-*overall <0.05), including BMI, history of sleep apnea, diabetes, with the highest values observed in cluster B3. Most echocardiographic parameters were comparable across clusters; however, right atrial volume index (RAVI) reflecting right atrial remodeling, was higher in cluster B1, which also had the lowest proportion of patients taking statins. Cluster B2 displayed intermediate values for most clinical parameters but was characterized by a higher prevalence of atrial fibrillation and a greater use of anticoagulant therapy. No significant differences were found for age, sex, heart rate, blood pressure, echocardiographic parameters, hypertension and ischemic etiology.

Biochemical parameters differed markedly across phenogroups (**Table 2**). Cluster B1 was associated with higher levels of markers reflecting disease severity, namely cardiac necrosis and fibrosis (troponin T; FGF-23 and soluble ST-2), liver dysfunction and fibrosis (Fib-4 index; NAFLD score; hypoalbuminemia; AST/ALT), congestion (CA-125) and platelet indices (platelet count, mean platelet volume). In contrast, cluster B2 was characterized by higher eGFR, reflecting better renal function and HDL-cholesterol levels. Cluster B3 exhibited higher values for parameters reflecting metabolic syndrome (glucose, HbA1c, triglycerides, metabolic scores and insulin resistance index). Other parameters, such as NT-proBNP, hemoglobin, and inflammatory markers such as C-reactive protein (CRP) did not differ significantly between clusters. Targeted amino and organic acid profiling revealed only modest differences across clusters, with only serine levels being significantly higher in cluster B1 (**Supplementary Table S3**).

**Table 2.**
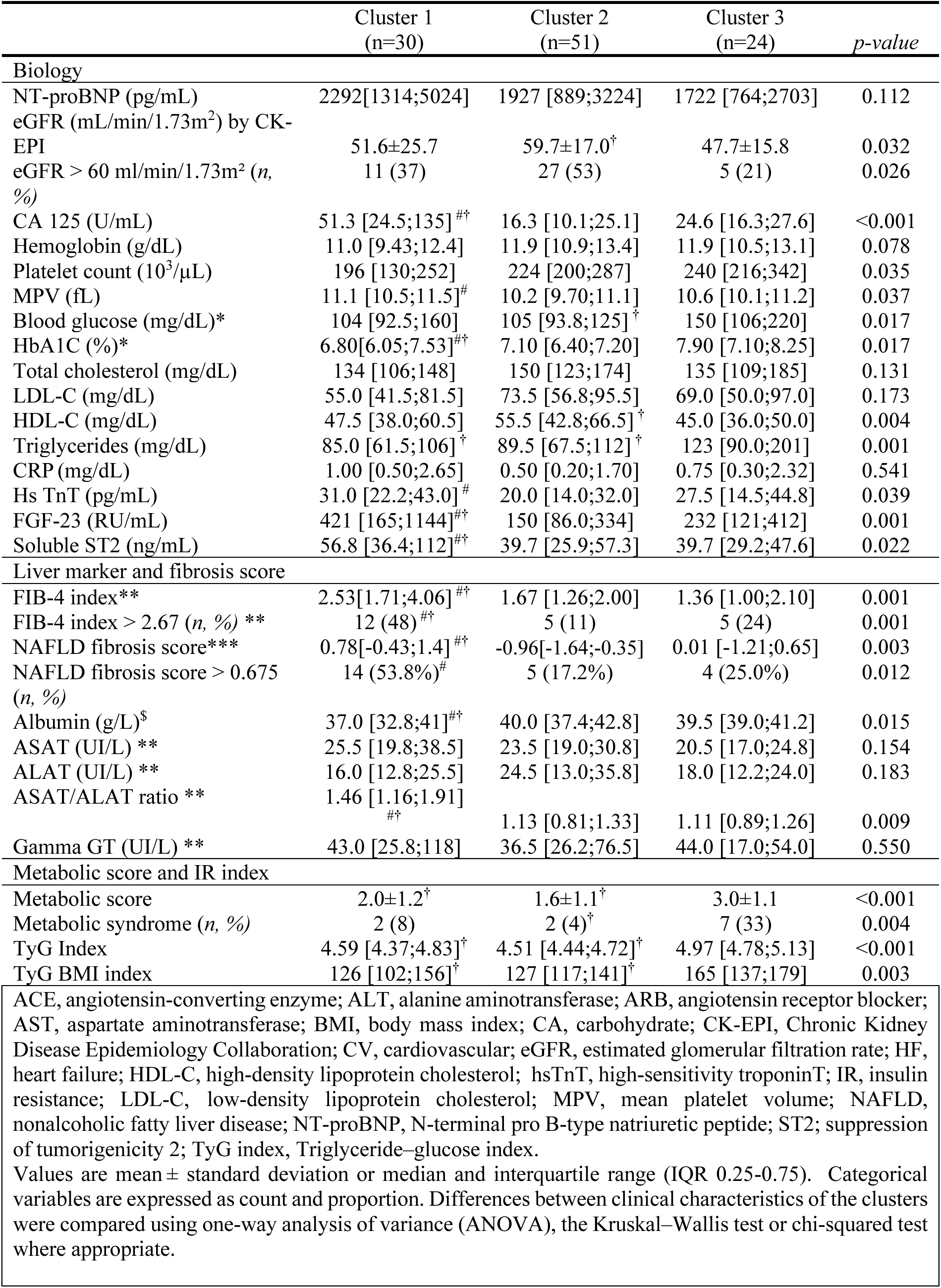
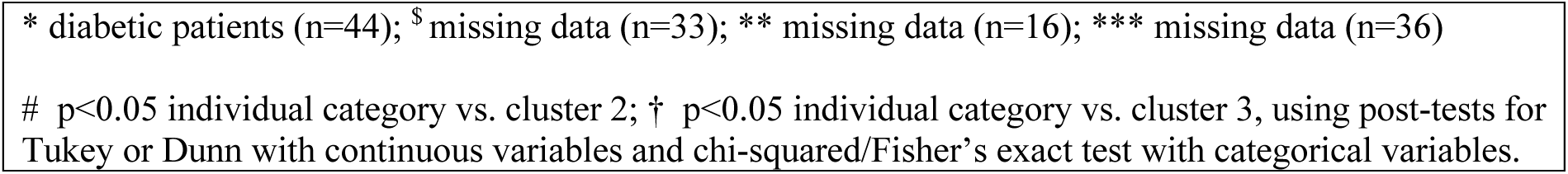
Baseline biochemical parameters of HFpEF patients according to cluster subgroup in the primary Belgian cohort.

|  | Cluster 1<br>(n=30) | Cluster 2<br>(n=51) | Cluster 3<br>(n=24) | *p-value* |
| --- | --- | --- | --- | --- |
| **Biology** |  |  |  |  |
| NT-proBNP (pg/mL) | 2292[1314;5024] | 1927 [889;3224] | 1722 [764;2703] | 0.112 |
| eGFR (mL/min/1.73m<sup>2</sup>) by CK-EPI | 51.6±25.7 | 59.7±17.0<sup>†</sup> | 47.7±15.8 | 0.032 |
| eGFR > 60 ml/min/1.73m<sup>2</sup> (n, %) | 11 (37) | 27 (53) | 5 (21) | 0.026 |
| CA 125 (U/mL) | 51.3 [24.5;135]<sup>#†</sup> | 16.3 [10.1;25.1] | 24.6 [16.3;27.6] | <0.001 |
| Hemoglobin (g/dL) | 11.0 [9.43;12.4] | 11.9 [10.9;13.4] | 11.9 [10.5;13.1] | 0.078 |
| Platelet count (10<sup>3</sup>/μL) | 196 [130;252] | 224 [200;287] | 240 [216;342] | 0.035 |
| MPV (fL) | 11.1 [10.5;11.5]<sup>#</sup> | 10.2 [9.70;11.1] | 10.6 [10.1;11.2] | 0.037 |
| Blood glucose (mg/dL)* | 104 [92.5;160] | 105 [93.8;125]<sup>†</sup> | 150 [106;220] | 0.017 |
| HbA1C (%)* | 6.80[6.05;7.53]<sup>#†</sup> | 7.10 [6.40;7.20] | 7.90 [7.10;8.25] | 0.017 |
| Total cholesterol (mg/dL) | 134 [106;148] | 150 [123;174] | 135 [109;185] | 0.131 |
| LDL-C (mg/dL) | 55.0 [41.5;81.5] | 73.5 [56.8;95.5] | 69.0 [50.0;97.0] | 0.173 |
| HDL-C (mg/dL) | 47.5 [38.0;60.5] | 55.5 [42.8;66.5]<sup>†</sup> | 45.0 [36.0;50.0] | 0.004 |
| Triglycerides (mg/dL) | 85.0 [61.5;106]<sup>†</sup> | 89.5 [67.5;112]<sup>†</sup> | 123 [90.0;201] | 0.001 |
| CRP (mg/dL) | 1.00 [0.50;2.65] | 0.50 [0.20;1.70] | 0.75 [0.30;2.32] | 0.541 |
| Hs TnT (pg/mL) | 31.0 [22.2;43.0]<sup>#</sup> | 20.0 [14.0;32.0] | 27.5 [14.5;44.8] | 0.039 |
| FGF-23 (RU/mL) | 421 [165;1144]<sup>#†</sup> | 150 [86.0;334] | 232 [121;412] | 0.001 |
| Soluble ST2 (ng/mL) | 56.8 [36.4;112]<sup>#†</sup> | 39.7 [25.9;57.3] | 39.7 [29.2;47.6] | 0.022 |
| **Liver marker and fibrosis score** |  |  |  |  |
| FIB-4 index** | 2.53[1.71;4.06]<sup>#†</sup> | 1.67 [1.26;2.00] | 1.36 [1.00;2.10] | 0.001 |
| FIB-4 index > 2.67 (n, %) ** | 12 (48)<sup>#†</sup> | 5 (11) | 5 (24) | 0.001 |
| NAFLD fibrosis score*** | 0.78[-0.43;1.4]<sup>#†</sup> | -0.96[-1.64;-0.35] | 0.01 [-1.21;0.65] | 0.003 |
| NAFLD fibrosis score > 0.675 (n, %) | 14 (53.8%)<sup>#</sup> | 5 (17.2%) | 4 (25.0%) | 0.012 |
| Albumin (g/L)<sup>\$</sup> | 37.0 [32.8;41]<sup>#†</sup> | 40.0 [37.4;42.8] | 39.5 [39.0;41.2] | 0.015 |
| ASAT (UI/L) ** | 25.5 [19.8;38.5] | 23.5 [19.0;30.8] | 20.5 [17.0;24.8] | 0.154 |
| ALAT (UI/L) ** | 16.0 [12.8;25.5] | 24.5 [13.0;35.8] | 18.0 [12.2;24.0] | 0.183 |
| ASAT/ALAT ratio ** | 1.46 [1.16;1.91]<sup>#†</sup> | 1.13 [0.81;1.33] | 1.11 [0.89;1.26] | 0.009 |
| Gamma GT (UI/L) ** | 43.0 [25.8;118] | 36.5 [26.2;76.5] | 44.0 [17.0;54.0] | 0.550 |
| **Metabolic score and IR index** |  |  |  |  |
| Metabolic score | 2.0±1.2<sup>†</sup> | 1.6±1.1<sup>†</sup> | 3.0±1.1 | <0.001 |
| Metabolic syndrome (n, %) | 2 (8) | 2 (4)<sup>†</sup> | 7 (33) | 0.004 |
| TyG Index | 4.59 [4.37;4.83]<sup>†</sup> | 4.51 [4.44;4.72]<sup>†</sup> | 4.97 [4.78;5.13] | <0.001 |
| TyG BMI index | 126 [102;156]<sup>†</sup> | 127 [117;141]<sup>†</sup> | 165 [137;179] | 0.003 |

ACE, angiotensin-converting enzyme; ALT, alanine aminotransferase; ARB, angiotensin receptor blocker; AST, aspartate aminotransferase; BMI, body mass index; CA, carbohydrate; CK-EPI, Chronic Kidney Disease Epidemiology Collaboration; CV, cardiovascular; eGFR, estimated glomerular filtration rate; HF, heart failure; HDL-C, high-density lipoprotein cholesterol; hsTnT, high-sensitivity troponinT; IR, insulin resistance; LDL-C, low-density lipoprotein cholesterol; MPV, mean platelet volume; NAFLD, nonalcoholic fatty liver disease; NT-proBNP, N-terminal pro B-type natriuretic peptide; ST2; suppression of tumorigenicity 2; TyG index, Triglyceride–glucose index.
Values are mean ± standard deviation or median and interquartile range (IQR 0.25-0.75). Categorical variables are expressed as count and proportion. Differences between clinical characteristics of the clusters were compared using one-way analysis of variance (ANOVA), the Kruskal–Wallis test or chi-squared test where appropriate.
\* diabetic patients (n=44); <sup>§</sup> missing data (n=33); \*\* missing data (n=16); \*\*\* missing data (n=36)

Leveraging the longitudinal follow-up data from the primary Belgian cohort, we tested the differential survival probabilities of the HFpEF patient clusters. Over a mean follow-up period of 22 [6-57] months, 44 patients (46%) died and 61 patients (58%) were hospitalized for HF, with 74 patients reaching the primary composite endpoint of all-cause mortality or HF hospitalization, whichever came first. Kaplan-Meier analyses stratified by phenogroups demonstrated significantly lower event-free survival (*p* = 0.0037) and overall survival (*p* = 0.00084) in cluster B1 compared with clusters B2 and B3 (**Figure 3**). In multivariable Cox models adjusted for established clinical, biochemical, and imaging covariates (NYHA classification 3-4, diabetes, chronic obstructive pulmonary disease, E/e’ ratio, hemoglobin, eGFR), cluster B1 remained associated with a higher risk of the composite endpoint (Hazard Ratio 1.98, 95% CI 1.19–3.82; p = 0.008; **Supplementary Table S4**). Altogether these findings underscore the ability of circulating lipids to identify HFpEF phenogroups with specific and clinically relevant characteristics and survival outcomes, with one subgroup characterized by a higher burden of adverse biomarkers and poorer prognosis.

**Figure 3:**
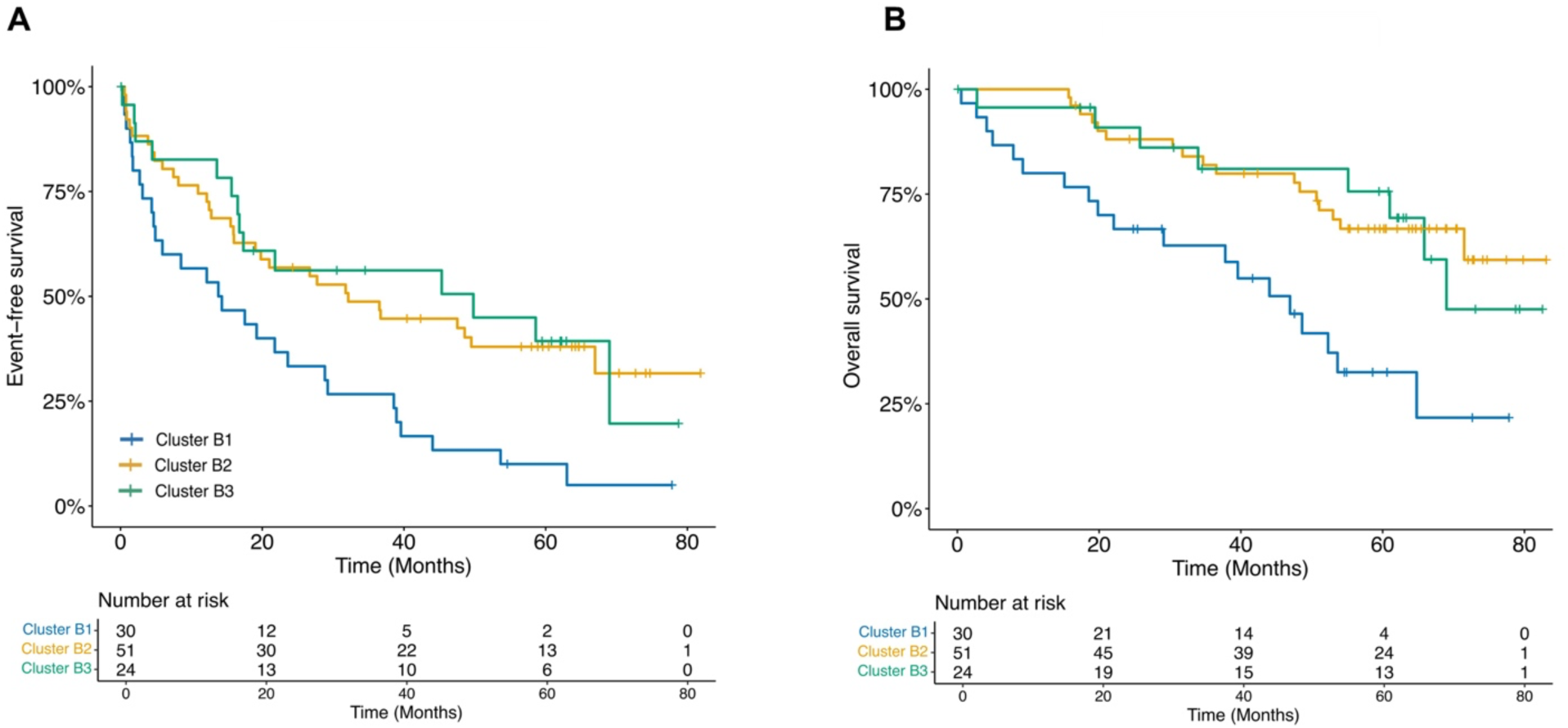
Differential survival of HFpEF phenogroups in the Belgian cohort. Kaplan–Meier survival curves of HFpEF patients stratified by lipidomic phenogroup (B1–B3) in the primary Belgian cohort. **(A)** Event-free survival for the composite endpoint of all-cause mortality or heart-failure hospitalization. **(B)** Overall survival for all-cause mortality. Unadjusted analyses show significantly poorer outcomes for cluster B1 compared with clusters B2 and B3 for event-free (*p* = 0.0037) and overall (*p* = 0.00084) survival. Numbers at risk are indicated below each plot.

### Cross-cohort comparison of lipidomic HFpEF phenogroups

To evaluate the extent to which lipidomic HFpEF phenogroups identified in the Belgian cohort are recapitulated in an independent population, we analyzed plasma lipidomic profiles from a secondary Canadian cohort. This cohort includes non-HF subjects (n=103) and HFpEF patients (n=74) from the MHI Biobank with baseline characteristics that differed from the primary Belgian cohort: HFpEF patients in the Canadian cohort had a higher BMI and a greater proportion of males, while non-HF subjects had more CV risk factors (**Supplementary Table S1** vs. Table 1 from Pouleur et al. 2024^31^). Despite these differences, similar plasma amino and organic acid levels were observed across cohorts, with only marginal differences between HFpEF patients and non-HF subjects in the Canadian cohort (**Supplementary Tables S5**).

Using the 158 annotated lipids common to both datasets (Supplementary Excel File) to enable direct cross-cohort comparison, unsupervised hierarchical clustering of HFpEF patients in the Canadian cohort also identified three lipidomic clusters (C1–C3; Supplementary Text). These clusters were associated with distinct baseline biochemical and metabolite profiles (**Supplementary Tables S6,7**; Supplementary Text). Visualization of HFpEF clusters in both cohorts using PCA illustrates differences in the structure of lipidomic variability between cohorts (**Supplementary Figure S6**). Several HFpEF clusters exhibited similar lipid patterns across cohorts, whereas others diverged (**Figure 4A**), based on lipidomic cluster signatures: C1 and C2 were most similar to cluster B3 from the Belgian discovery cohort, while cluster C3 aligned most closely with cluster B2. These shared profiles clustered nearer to non-HF subjects from both cohorts than to the high-risk B1 cluster. No cluster in the validation cohort exhibited a lipidomic signature comparable to B1, which displayed the most distinct lipid patterns.

**Figure 4.**
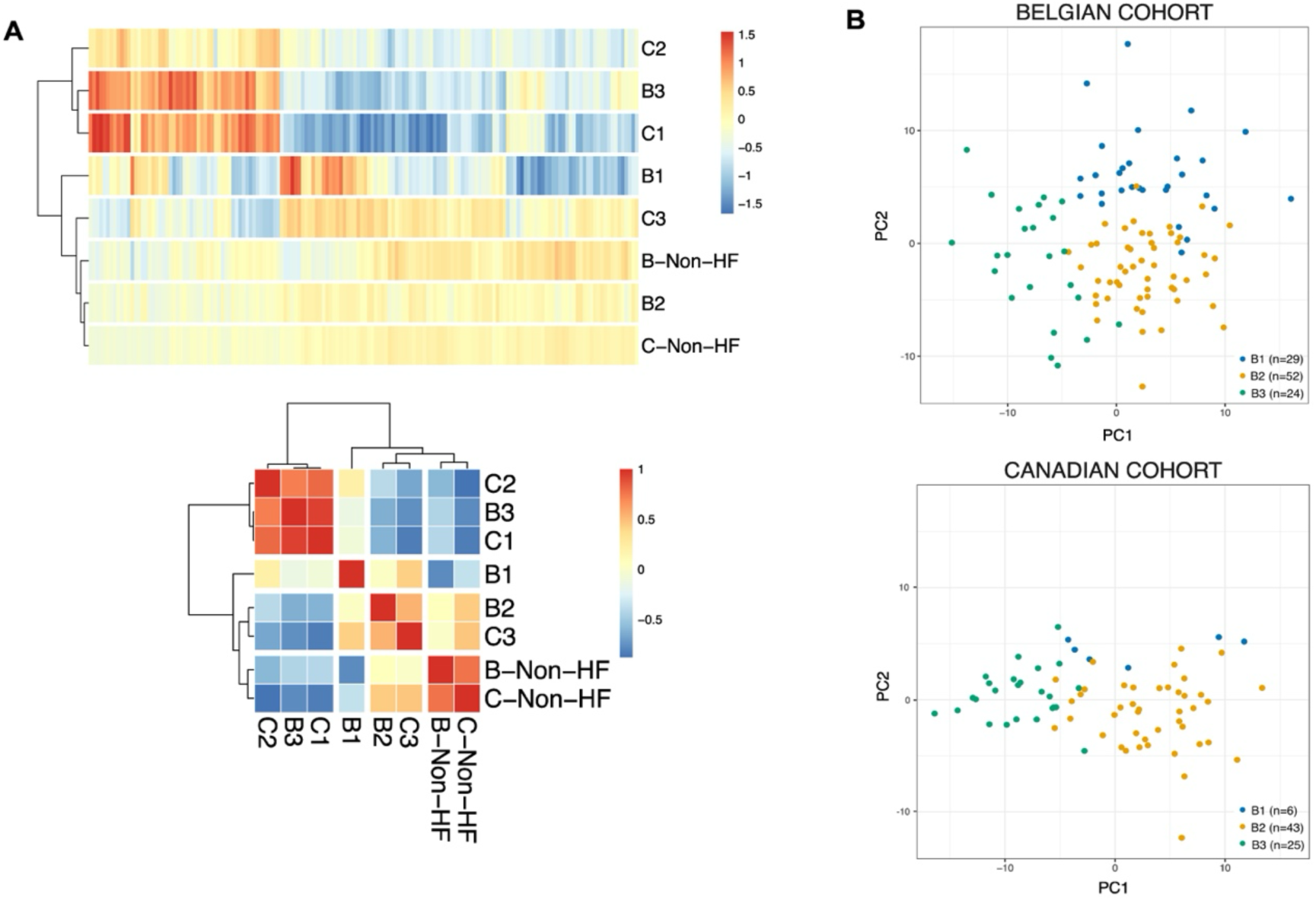
Cross-cohort comparison of lipidomic HFpEF phenogroups between Belgian and Canadian cohorts. **(A)** Heatmaps comparing global lipidomic signatures of HFpEF clusters identified in the Belgian (B1-B3) and Canadian (C1-C3) cohorts using the 158 annotated lipids shared between datasets. The upper panel shows relative mean lipid abundance across individual subjects from each cluster and non-HF groups, while the lower panel displays the Pearson correlations between these groups, highlighting similarities and divergences in lipid subclass patterns across cohorts. **(B)** Cross-cohort projection of HFpEF patients using a classification model trained on Belgian clusters. PCA score plots show Belgian cohort patients (upper panel) and Canadian cohort patients (lower panel) represented in a shared lipidomic feature space based on the common lipid set. Most Canadian cohort patients map to B2- or B3-like regions, whereas few exhibit a B1-like profile, indicating limited representation of the high-risk lipidomic phenogroup in the Canadian cohort. Numbers of subjects per cluster are indicated in parentheses.

To further assess cross-cohort correspondence at the patient level, a random forest classifier trained on Belgian HFpEF clusters using the shared lipid set was applied to the HFpEF patients of the Canadian cohort (**Figure 4B**). While most BECAME-HF2 patients were classified as B2- or B3-like, only a small fraction (8%) was predicted as B1-like (**Supplementary Figure S7**), indicating limited representation of this lipidomic phenogroup in the Canadian cohort. The high-risk lipid signature identified in Belgian cluster B1 was underrepresented in the Canadian cohort, motivating further investigation of this patient population in the Canadian disease context.

### Minimal lipid signature of HFpEF cluster with poor prognosis

Considering that HFpEF cluster B1 shows the worst event-free survival prognosis and the most adverse clinical profile (**Figure 3**; **Tables 1-2**), we sought to identify a minimal lipidomic signature associated with this high-risk subgroup and to explore whether it could help recognize similar patient profiles, independent of cohort composition. Using the 235 annotated lipids in BECAME-HF1, a regularized feature-selection strategy was applied (**Methods**) and identified a parsimonious set of 10 lipids spanning six lipid subclasses with discriminative capacity (**Supplementary Figure S8A**). These 10 lipids occupy central positions within the global lipid correlation network, mapping onto the major lipidomic structure observed across HFpEF-associated lipids (**Figure 5A**; **Supplementary Figure S8B**). Differences of lipid levels between clusters for these 10 lipids (**Figure 5B**) highlight that B1 patients not only differ from clusters B2 and B3, but also from non-HF subjects. The signature is characterized by higher levels of acylcarnitines (CAR 18:1, CAR 26:1) along with one oxidized SM (SM(d18:0/16:0(O)) and two ether PC (PC(0-18:1/16:0) and (PC(O-20:2/20:4), and lower levels for two PCs, one ether PE (PE(O-16:0/22:6), one TG, all with a PUFA acyl chain (20:4 or 22:6), as well as one SM with an odd VLCFA.

**Figure 5.**
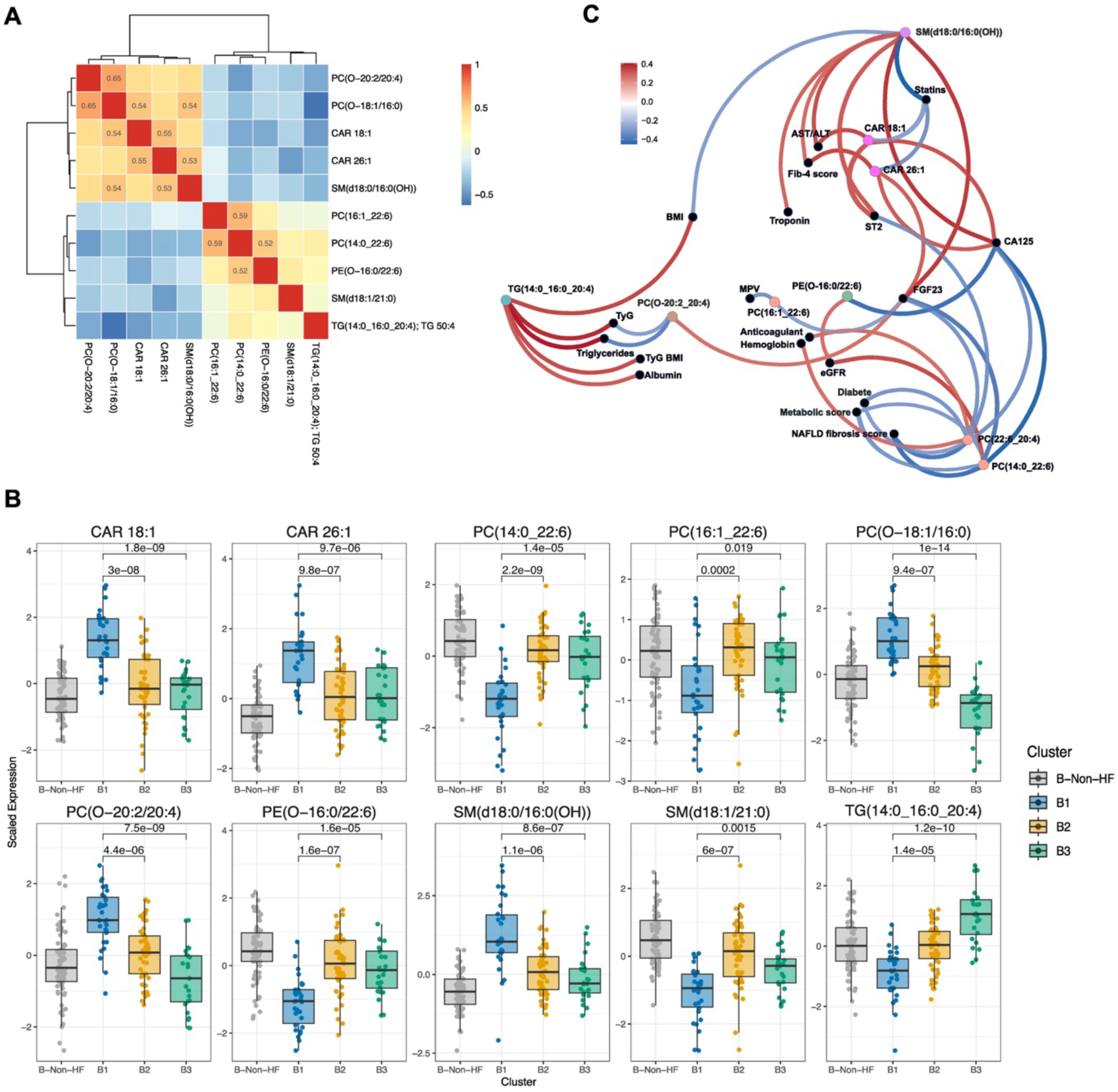
Minimal lipid signature identifying the high-risk HFpEF phenogroup and its clinical correlates in the Belgian cohort. **(A)** Correlation matrix of the 10 lipids comprising the minimal signature selected by regularized feature selection (LASSO) with cross-validation from the 235 annotated lipids. These lipids occupy central positions within the global lipid correlation network and represent multiple lipid subclasses. **(B)** Box plots showing scaled plasma levels of the signature lipids across HFpEF phenogroups (B1-B3) and non-HF subjects. The high-risk B1 cluster exhibits a distinct lipid profile characterized by increased acylcarnitines, oxidized sphingomyelins, and ether phosphatidylcholines, and decreased polyunsaturated phosphatidylcholines, ether phosphatidylethanolamines, triglycerides, and very-long-chain sphingomyelins. **(C)** Correlation network linking signature lipids with clinical and biochemical parameters in HFpEF patients. Most signature lipids are significantly associated with markers of disease severity, including cardiac injury and fibrosis (TnT, FGF-23, sST2), hepatic dysfunction and fibrosis (Fib-4 index, NAFLD score), and congestion (CA-125), supporting their relevance to the adverse cardiometabolic phenotype of cluster B1.

Most of these lipids were significantly associated with clinical and biochemical markers of disease severity among HFpEF patients (**Figure 5C**; **Supplementary Figure S9**). Notably, the majority are associated with markers of disease severity reflecting cardiac and liver necrosis or fibrosis (TnT, FGF-23, sST2, Fib-4, NAFLD score) and congestion (CA-125), which are all distinctive clinical characteristics of cluster B1. Thus, this analysis supports their relevance to the high-risk HFpEF phenotype represented by cluster B1 and underlines its potential utility for identifying patients with similar adverse cardiometabolic profiles.

## 5. Discussion

HFpEF is a complex heterogeneous syndrome for which patient stratification remains a major barrier to mechanistic understanding and therapeutic progress. In this study, we demonstrate that comprehensive untargeted plasma lipidomics captures previously unrecognized metabolic heterogeneity within HFpEF and enables clinically meaningful phenogrouping across cohorts. In a deeply phenotyped real-world Belgian cohort with longitudinal outcomes, we identified three lipid-driven distinct HFpEF phenogroups associated with divergent clinical profiles and prognosis, including a high-risk subgroup associated with worse adverse outcomes. This subgroup was underrepresented in the secondary Canadian cohort, conveying patient heterogeneity across cohorts. Additionally, we identified a minimal lipid signature that delineates the high-risk HFpEF phenogroup, providing a focused molecular framework to support future mechanistic studies and refined phenotyping of this underrecognized HFpEF population.

Recent efforts to enhance the mechanistic insight in HFpEF phenomapping have leveraged machine-learning approaches applied to high-dimensional omics data, particularly plasma proteomics, thereby uncovering patient subgroups with distinct biology and clinical outcomes (e.g., Sanders-van Wijk *et al.*^20^). To date, the use of plasma targeted metabolomics^37^ showed minimal discriminatory power to distinguish HFpEF phenogroups, supporting our findings with targeted profiling of amino and organic acids. However, our findings demonstrate the unique capacity of comprehensive untargeted lipidomics in revealing multiple lipid perturbations in HFpEF patients, which are consistent with proposed metabolic mechanisms, namely mitochondrial and peroxisomal dysfunction, substrate overload, oxidative stress, and sphingolipid acyl-chain remodeling linked to altered cell signaling, while pointing to novel ones, as discussed in detail below. Of note, our findings contrast with those of Jovanovic *et al.*^38^ of marginal lipid associations with HFpEF, likely explained by differences in cohort size, analytical design, and lipidomic methodology. Our extended chromatographic separation likely enabled detection of lipid isomers and subtle subclass-specific changes that may be obscured by shotgun approaches. Importantly, we have shown that data analysis based on case-control comparison obscures meaningful heterogeneity, as several lipid subclasses exhibit opposing directions of change across HFpEF phenogroups.

The lipid-driven HFpEF phenogroups identified in the Belgian cohort were associated with distinct clinical and biochemical profiles as well as long-term survival outcomes, supporting their clinical relevance beyond molecular separation alone. HFpEF patients in cluster B3 displayed hallmark features of the well-described obese-metabolic HFpEF phenotype^13,14^, characterized by metabolic syndrome features with obesity associated with substrate overload (high plasma glucose and TG; Table 1-2). In contrast, clusters B1 and B2 could not be readily mapped onto described HFpEF phenotypes. Notably, they did not differ across several parameters commonly used for HFpEF stratification^14^(such as age, NT-proBNP, CRP, most echocardiographic indices and traditional cardiovascular risk factors) highlighting the limited discriminative power of standard clinical variables for resolving HFpEF biological heterogeneity.

Cluster B1 exhibited the worst prognosis and was distinguished by a constellation of biomarkers indicative of advanced multisystem involvement, including myocardial injury and fibrosis, congestion, platelet activation, and notably, liver dysfunction. Elevated fibrosis indices, higher NAFLD scores, and lower albumin levels point to a prominent cardio-hepatic axis that remains underexplored in HFpEF, despite reports of similar patient profiles in other cohorts (TOPCAT cohort^15^. HFpEF and liver diseases such as metabolic dysfunction–associated fatty liver disease (MAFLD, previously referred to as NAFLD) share major pathogenic risk factors and events, which include diabetes and lipid metabolic abnormalities. On this basis, it was proposed that “MAFLD-related HFpEF” should be recognized as a distinct phenotype^10,39^, although it remains to be ascertained whether liver disease precedes or is a consequence of HFpEF. Importantly, the metabolic profile of cluster B1 differed from that of the obese–metabolic cluster B3, lacking overt obesity or insulin resistance, yet displaying a comparable prevalence of diabetes, which together with lower albumin levels, is consistent with a frailty- or sarcopenia-associated metabolic state, which may contribute to their particularly poor prognosis.

Cluster B2 was most consistent with an aging-related HFpEF phenotype, having a higher prevalence of atrial fibrillation but otherwise favorable clinical and biochemical characteristics, including preserved renal function, higher HDL-cholesterol levels, lower markers of disease severity, and better survival outcomes (**Table 2**, **Figure 3**). These features distinguish cluster B2 from the advanced (B1) and obese-metabolic (B3) phenotypes and support the notion that aging-related cardiac remodeling represents a distinct axis of HFpEF heterogeneity.

To explore mechanisms underlying lipid-defined HFpEF heterogeneity, we will focus on the high-risk cluster B1 and the minimal lipid signature that discriminates this subgroup (**Figure 6**), indicated thereafter in the text by an asterisk (*Lipid). We will also consider additional subsets of lipids distinguishing individual clusters from the others (**Supplementary Figure S5)**. The minimal lipid signature correlates with clinical and biochemical markers of disease severity and implicates multiple metabolic pathway dysfunctions or pathogenic mechanisms. Consistent with prior HF studies, we found (i) increase in several LC-CARs (particularly *CAR 18:1), indicative of mitochondrial β-oxidation defects and defective energy metabolism^22,23,40^, (ii) increased VLC-CAR (*CAR 26:1), a marker of peroxisomal metabolic dysfunction^41^, and (iii) reduced VLCFA-containing SM (*SM(d18:1/21:0)), reflecting altered cell-death signaling pathways^42–44^. In addition, we observed previously unreported changes in PUFA-containing PC (reduced; *PC(14:0_22:6); *PC(16:1_22:6); TG (reduced; *TG(14:0_16:0_20:4); as well as ether PL, both PC (increased; *PC(O-20:2/20:4 *PC(O-16:0/22:6)) and PE (reduced; *PE(16:0_22:6), and finally, one oxidized SL (increased; *SM(d18:0/16:0(OH)).

**Figure 6.**
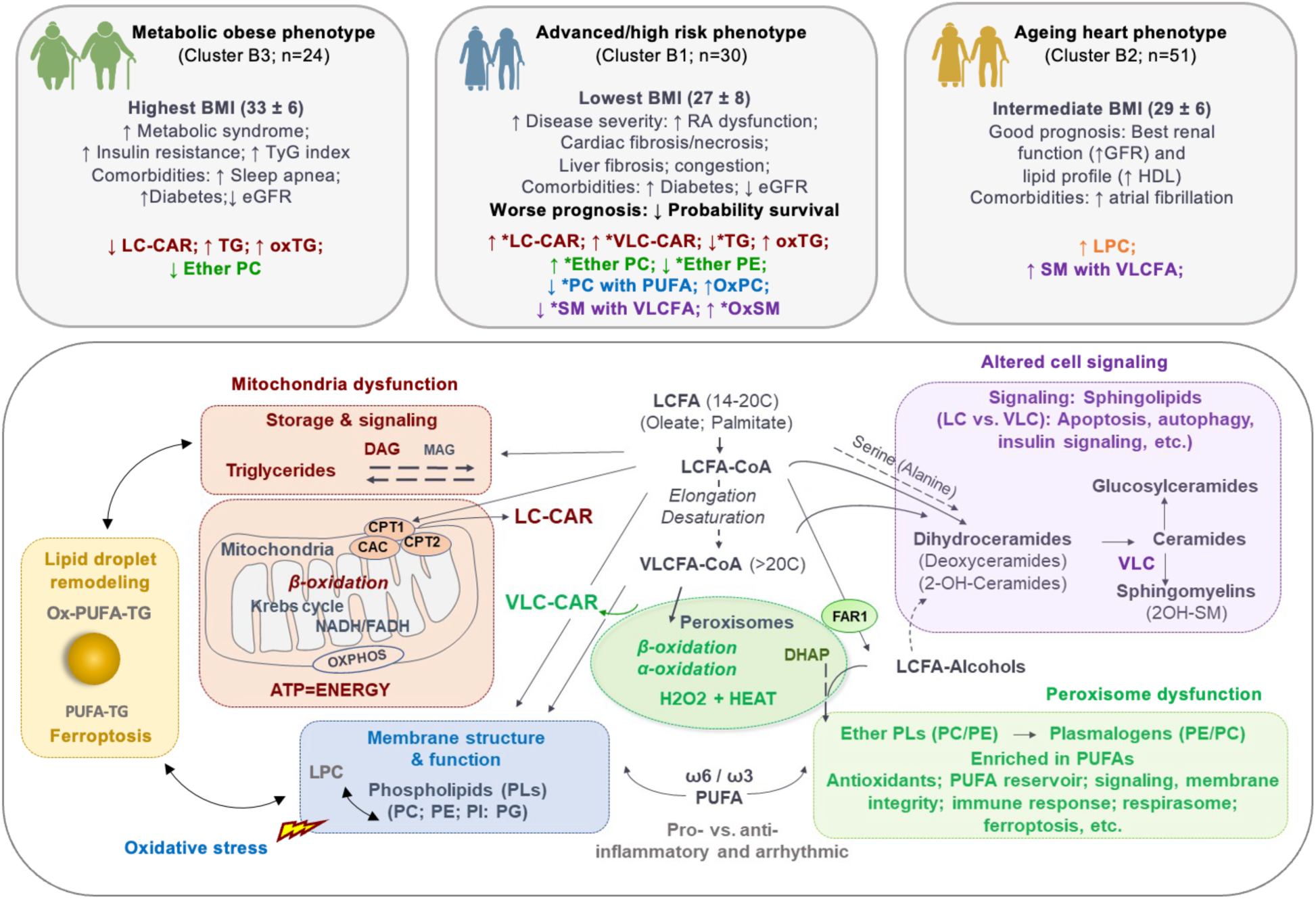
Proposed mechanisms underlying lipid-defined HFpEF phenogroups. Schematic overview of major metabolic pathways of long-chain fatty acids (LCFA) and their potential contribution to the plasma lipid signatures distinguishing HFpEF phenogroups. Color coding highlights functional domains: mitochondria-related energy metabolism and lipid storage (red), peroxisome-related metabolism (green), sphingolipid signaling (purple), and the oxidative stress–sensitive processes membrane phospholipid metabolism (blue) and lipid droplets remodeling (yellow). Major clinical characteristics discriminating the three HFpEF clusters are summarized along with lipid subclasses that distinguish individual clusters from the others (Fig. 5B; Fig. S5). Subclasses that are part of the high-risk minimal lipid signature (Fig. 5B) are indicated by an asterisk (*Lipid). The high-risk B1 cluster is characterized by lipid alterations consistent with mitochondrial dysfunction, impaired peroxisomal metabolism, altered sphingolipid signaling as well as oxidative stress-induced alterations in membrane phospholipid composition and lipid droplet remodeling, potentially contributing to cardiac and extra-cardiac pathology including liver dysfunction.

Elevated plasma levels of *CAR 18:1 and *CAR 26:1 correlate positively with several markers of disease severity (**Figure 5C**, **Supplementary Figure S9**), consistent with prior association with adverse prognosis^26^, and potentially pro-inflammatory or pro-arrhythmogenic effects^40^. Although plasma levels of *CAR 18:1 were found to be higher in HFrEF than HFpEF ^22,23^, our results show heterogeneity within HFpEF, with highest levels in the high-risk B1 cluster, including relative to the obese-metabolic B3 phenotype. Although the exact tissue sources of these LC- and VLC-CAR cannot be determined, our results suggest an extra-cardiac origin, potentially from liver^45^, consistent with the elevated liver disease markers observed in B1.

Peroxisomal dysfunction is further supported by altered plasma levels of ether PLs, precursors of plasmalogens^41^. These are major cell membranes components contributing to antioxidant defense and cellular signaling, notably ferroptosis^46^, with both processes implicated in HFpEF pathogenesis. Notably, ether PC (increased) and ether PE (reduced) showed opposite changes; these lipid classes are enriched in cardiac and hepatic tissues respectively^47^, suggesting tissue-specific dysregulated metabolism that may influence ferroptosis. In cells, including cardiomyocytes, ferroptosis was found to be regulated by PUFA-containing ether PL, whereby their downregulation confers resistance and upregulation sensitivity^46^. Reduced PUFA-containing PCs and increased oxidized PC (PC(16:0_18:2(OH)); **Figure S5A**) in B1 indicate enhanced oxidative stress. Although both B1 and B3 exhibited elevated oxidized TG (TG(16:0_18:1_18:3(O))) relative to B2 (**Figure S5B**), cluster B3 showed opposite changes in PUFA-containing ether PC, PC and TG species compared to B1. We speculate that this pattern reflects differential lipid droplet remodeling under conditions of oxidative stress, a process proposed to mitigate susceptibility to cell death by ferroptosis by sequestering PUFA as well as oxidized PUFA away from membranes PL^48,49^. The markedly higher oxidized PC levels in B1 suggests that this protective buffering mechanism may be overwhelmed in this cluster, whereas it can be sustained in B3 by the greater TG availability associated with obesity and insulin resistance.

Cluster-specific alterations in SL, particularly the SM subclass, further support differential risk profiles. Elevated plasma levels of SLs containing LCFA (16 to 18 carbons) vs VLCFA (>20 carbons) have been associated with poorer prognosis in cardiovascular disease^28,50–52^. In our study, VLCFA-containing SMs (*SM(d18:1/21:0) were lowest in cluster B1 and highest in cluster B2, which may contribute to more favorable prognosis. Of note, cluster B2 had highest plasma levels for lysophosphatidylcholine (LPC) species (Figure S5), whose circulating and/or myocardial levels were reported to be altered in sepsis^53^ and cancer-related cardiotoxicity^54^. More importantly, their serum levels inversely predicted increased mortality in patients with acute HF^55^. Finally, interestingly, B1 showed the highest plasma level of oxidized SM (*SM(d18:0/16:0(OH)), which showed positive correlation with disease markers similarly to *CAR 18:1 and *CAR 26:1. The oxidized SM species is likely to be a 2-hydroxylated SM generated by the fatty acid 2-hydroxylase (FA2H), whose expression level has been implicated in obesity-induced insulin resistance, neurodegeneration and tumor progression^56,57^. The role of these lipids in HFpEF remains to be clarified.

Several aspects of this study highlight both limitations and opportunities for future work. While observational and cross-sectional by design, the integration of deep clinical phenotyping with untargeted lipidomics enabled the identification of robust, clinically meaningful HFpEF subgroups and provides a strong foundation for hypothesis generation and translational follow-up. Although the overall sample size was modest, it remains larger than most metabolomic/lipidomic studies in cardiovascular disease. The identification of a high-risk HFpEF phenogroup despite its relative underrepresentation in the Canadian cohort underscores the need for targeted detection strategies. In this context, the derivation of a minimal lipid signature represents an important step toward the development of clinically scalable tools to identify such patients. Because many lipid species are highly correlated in plasma lipidomics data, the lipids retained in the signature likely represent broader lipid modules rather than uniquely causal biomarkers. From a technical perspective, the inclusion of partially annotated features reflects the breadth of untargeted lipidomics and highlights opportunities for future structural characterization and pathway refinement as lipid databases evolve. Independent validation in larger cohorts will be required to confirm the robustness and generalizability of this lipidomic signature.

An important consideration is that patients in both cohorts were recruited prior to the widespread use of SGLT2 inhibitors in HFpEF. While this limits direct inference regarding treatment-modified phenotypes, it also reduces therapeutic confounding. Given prior evidence of lipidomic modulation in response to SGLT2 inhibition^29^, it will be important in future studies to test whether lipidomic phenogroups exhibit differential responses to SGLT2 inhibition in HFpEF populations. Finally, differences in cohort composition emphasize the importance of multi-population validation, while the integration of direct and longitudinal measures of liver fibrosis, skeletal muscle mass, and congestion represents a clear opportunity to further refine and mechanistically anchor the proposed cardio-hepatic HFpEF phenotype. More broadly, these findings highlight the potential of integrating lipidomics with complementary omics layers, imaging, and clinical data in large, well-powered cohorts. Such integrative approaches can help uncover the biological mechanisms underlying distinct HFpEF subgroups and ultimately enable more precise, mechanism-guided therapeutic strategies.

## Data Availability

The lipidomic and metabolomic datasets generated and analyzed in this study are derived from human clinical cohorts. Processed lipidomic data matrices (log₂-transformed, normalized, and batch-corrected), including lipid annotations and cluster assignments, are available in a public repository (to be provided). Summary-level data supporting the main findings of this study are provided in the Supplementary Tables. All custom code used for lipidomic data processing, statistical analyses, clustering, and visualization was written in R and is available on GitHub [https://github.com/HussinLab/BECAME_HF/] with appropriate documentation. Any additional information required to reanalyze the raw data is available from the corresponding authors upon reasonable request, subject to institutional data-sharing agreements and ethical approval.

## Funding

BECAME-HF was supported by bilateral research programs between the Fonds de Recherche du Quebec (FRQ, Québec, Canada) and the Fonds National de la Recherche Scientifique (FNRS, Brussels, Belgium) (Bilateral research projects FRQ-FNRS - PINT-BILAT-P 2018), by the Action de Recherche Concertée of the Wallonia-Brussels Federation (ARC 23/28-132), and by grants from the “Fondation Saint-Luc” (Brussels, Belgium). This project also benefited from infrastructure supported by the Canada Foundation for Innovation (grant numbers 12126, 20415, and 36283 to JCT and CDR) and the MHI Foundation. Computational work was funded in part by a National Sciences and Engineering Research Council (NSERC) Discovery Grant to JGH (RGPIN-2022-04262). NM, SL and CR were supported by Fondation Saint Luc, Belgium. JGH holds a Tier 2 Canada Research Chair (CRC) in responsible multi-omics data science, JCT holds a Tier 1 CRC in personalized and translational medicine, ACP is post-doctorate Clinical Master Specialist at FNRS, SH is a senior research associate of FNRS.

## Acknowledgments

We thank Jean-Christophe Grenier and Douae Maatougui for valuable discussions regarding the computational analysis, as well as Caroline Daneault and Bertrand Bouchard for the lipidomic analysis. This work was made possible through computational resources provided by the Digital Research Alliance of Canada.

## Author contributions

JGH, SH, LB, CB, ACP, CDR conceived the study and secured funding. JGH, NM, SH, LB, CB, ACP, CDR supervised the project. NM, CR, SL, DV, DR, JCT, ACP, CB conducted the clinical study, including patient recruitment and phenotyping. AF and IFR performed metabolomic experiments and data acquisition. NM, PM, OT, JTL, GBr conducted computational and statistical analyses. JGH and GBo advised on statistical analyses. JGH, NM, PM, JTL, ACP, CB, CDR drafted the manuscript. JGH, NM, AF, PM, CDR prepared figures. All authors revised the manuscript and approved the final version.

## Declaration of Use of Artificial Intelligence (AI)-assisted technologies

During the preparation of this work the authors used Large Language Models to improve the readability and language of sections of the manuscript as well as for code optimization. After using these tools, the authors reviewed and edited the content as needed and take full responsibility for the content of the published article. No AI tools were used for interpretation or generation of scientific conclusions.

## Notes

### Competing Interest Statement

The authors have declared no competing interest.

### Author Declarations

The study was conducted in accordance with the Declaration of Helsinki and approved by local ethics committees in Belgium (Comite déthique Hospitalo-Facultaire, Cliniques universitaires Saint-Luc and UCLouvain; approvals 2012/23AVR/199 and 2018/19MAR/118) and Canada (Montreal Heart Institute protocol #2017-2316). All participants provided written informed consent.

