## Supplementary Material for "Lipidomics Identifies HFpEF Phenogroups and a High-Risk Metabolic Signature - The BElgian and CAnadian MEtabolomics in HFpEF (BECAME-HF) project"

The **BE**lgian and **CA**nadian **ME**tabolomics in **HF**pEF (BECAME-HF) project

*Running title: Lipid Phenogroups in HFpEF*

### Supplementary Material

#### Supplementary Text

##### 1. Cohorts and samples

This study includes subgroups of subjects from two cohorts, namely a primary Belgian (BECAME-HF1) cohort and a secondary Canadian cohort (BECAME-HF2), for which inclusion and exclusion criteria, as well as demographic, clinical and biochemical parameters were previously described in detail[1].

The real-life Belgian cohort is made of consecutive patients with HFpEF prospectively evaluated for inclusion in the study in a single center between December 2015 and June 2017. The present study reports data for 105 HFpEF patients and 72 age- and sex-matched non-HF subjects. The following criteria had to be fulfilled for study inclusion as HFpEF patients: New York Heart Association functional (NYHA) class ≥ II, typical symptoms and signs of HF, N-terminal pro-brain natriuretic peptide (NT-proBNP) >350 pg/mL and/or a hospitalization for HF in the previous 12 months, with preserved left ventricular ejection fraction LVEF (≥ 50%; HFpEF) and relevant structural heart disease [LV hypertrophy/left atrial (LA) enlargement] and/or diastolic dysfunction assessed by echocardiography for HFpEF, as previously described[2]. HFpEF patients were prospectively followed-up for clinical and survival status during ambulatory visits and/or phone calls at 6-month intervals. Clinical and survival status were obtained by follow-up visits and by phone contact with the patients, their relatives, or their physician. The primary endpoint was a composite of all-cause mortality or hospitalization for HF, whichever came first. Hospitalization was defined as patients treated in the emergency room or admitted to a hospital, diagnosed with decompensated HF, and requiring IV diuretics. The secondary endpoint was all-cause mortality. Non-HF subjects had no history of cardiovascular events/disease (including HF), no significant past medical history or chronic disease, but exhibited some cardiovascular risk factors. They were recruited by advertisement in the local community and underwent a full clinical examination, electrocardiogram, echocardiography, and exercise stress test, which all had to be normal prior to inclusion. All subjects underwent blood sampling and complete transthoracic echocardiography (iE33 system Philips). All echocardiographic measurements were averaged over three beats in atrial fibrillation.

The secondary Canadian cohort include participants from the Montreal Heart Institute (MHI) that were recruited between April and September 2018, including 74 HFpEF patients and 103 age- and sex-matched non-HF subjects. Statistical analysis for the comparisons of the two cohorts of subjects for demographic, clinical and biological parameters was previously described in detail[1]. These participants are part of the MHI Biobank, which includes over 17,000 participants of which 987 suffered from HF at baseline, and the study has been approved at the MHI under project MIRACLE #17-2306.

Blood samples were obtained by venipuncture at inclusion. After centrifugation at 3,500 rpm for 10 min, serum aliquots were stored at −80 °C for subsequent analysis of the following biomarkers, as previously described[1]. NT-proBNP and high-sensitive troponin T (hsTnT) levels were also measured with automated electrochemiluminescence immunoassay on the CobasR 8000 platform. Soluble ST2 and intact FGF-23 levels were determined by immunoassays. The present study includes also the following additional biomarkers, biological parameters and scores. CA 125 levels were determined with a two-sites automated electrochemiluminescence assay on the CobasR 8000 platform (Roche Diagnostics, Mannheim, Germany)[3]. The Fib-4 score is a non-invasive tool used to assess liver fibrosis risk in patients with chronic liver diseases like NAFLD. The formula for the Fib-4 score is: Age (in years) multiplied by AST (U/L), divided by the product of platelet count (PLT, 10^9/L) and the square root of ALT (U/L). A Fib-4 score greater than 2.67 indicates a high likelihood of advanced liver fibrosis. The NAFLD Fibrosis Score combines age, body mass index (BMI), impaired fasting glucose or diabetes status, AST/ALT ratio, platelet count, and albumin level. The formula is: -1.675 + (0.037 multiplied by age in years) + (0.094 multiplied by BMI) + (1.13 multiplied by impaired fasting glucose or diabetes status [1 if present, 0 if absent]) + (0.99 multiplied by the AST/ALT ratio) - (0.013 multiplied by platelet count [PLT, 10^9/L]) - (0.66 multiplied by albumin level in g/dL). A NAFLD Fibrosis Score greater than 0.675 indicates a high likelihood of advanced fibrosis. The Metabolic Syndrome is assessed when three or more of the following criteria are present: BMI >30, elevated triglycerides (≥150 mg/dL), low HDL cholesterol (<40 mg/dL for men, <50 mg/dL for women), high blood pressure (≥130/85 mmHg), and high fasting glucose (≥100 mg/dL). The Triglyceride-Glucose Index (TyG) is a marker of insulin resistance. It is calculated as the natural logarithm of the product of fasting triglycerides (mg/dL) and fasting glucose (mg/dL), divided by 2.

#### 2. Targeted Metabolomic analysis

All metabolomics analyses were performed between 2018 and 2021. Analyses were achieved on aliquots that were thawed once (Belgian cohort) or unthawed (Canadian cohort). Samples were processed on different batches, using a balanced design. Following recommended guidelines[38], stratified randomization of samples was achieved according to potential confounding factors, namely age, sex and disease status, to minimize potential batch effects.

Plasma levels of 21 amino acids and 12 organic acids were measured by isotope dilution gas chromatography-mass spectrometry (GC-MS) in a subset of the Belgian cohort (n=44 non-HF and n=70 HFpEF) and Canadian cohort (n=103 non-HF and n=74 HFpEF) as previously described[4,5] with slight modifications.

Plasma (100 µl) was extracted with methanol (70%) and hydroxylamine (100 µl, 1 M, pH 7.6). A mixture of internal and external standards of organic acids (100 nmol 13C-lactic acid, 30 nmol 13C-pyruvic acid, 15 nmol 13C4-β-hydroxyubutyric acid, 20 nmol d4-citrate, 20 nmol 13C4-ketoglutarate, 10 nmol d4-succinate, 10 nmol d3-malate, 10 nmol 13C4,d2-α-ketobutyric acid, and 60 nmol 13C4-acetoacetic acid), and a mixture of amino acids (70 nmol 13C3-alanine, 50 nmol 13C2-glycine, 35 nmol 13C5-valine, 25 nmol 13C6,15N-leucine, 10 nmol 13C6-isoleucine, 40 nmol 13C5,15N-proline, 5 nmol 13C5-methionine, 20 nmol 13C5,15N-serine, 25 nmol 13C4,15N-threonine, 10 nmol d5-phenylalanine, 2 nmol 13C4,15N-aspartate, 10 nmol 13C5,15N-glutamate, 15 nmol 13C6-arginine, 10 nmol 13C9-tyrosine, 10 nmol 13C4,15N2-asparagine, 80 nmol 13C5,15N2-glutamine, 40 nmol 13C6,15N2-lysine, 10 nmol 13C11,15N2-tryptophan, 150 nmol 13C3,15N cysteine and 20 nmol 13C6-histidine) were added to each sample. Samples were sonicated for 2 min and homogenized for 45 s with 2.8-mm zirconium oxide beads (Omni International). After addition of hydrochloric acid (50 µl, 1 M, pH between 5 and 6), samples were incubated at 70°C for 15 min and centrifuged at 22,000 g for 10 min. The supernatants were dried with Na2SO4, then evaporated and samples were solubilized in 25 µl pyridine at 45°C for 90 min. Derivatization was performed using 100 µL N-methyl-N-tertbutyldimethylsilyltrifluoroacetamide at 90°C for 4 h.

Samples (2 µl) were injected onto a gas chromatography-mass spectrometer (GC-MS; Agilent 6890N chromatograph coupled to a 5975N mass spectrometer) and operated in electronic ionization mode with helium as reagent gas at a flow rate maintained throughout [organic acids (OA): 23.3 ml/min and amino acids (AA):20.1 ml/min] in split mode (OA; 28:1 and AA; 23.2:1). The temperature program for OA was fixed as follows: 150°C for 5 min, increment of 10°C/min up to 300°C, and then 20°C/min up to 320°C. For AA, the temperature program was 150°C for 3 min, increment of 7°C/min up to 210°C and maintained constant during 3 min, increment of 7°C/min up to 310°C and maintained constant during 3 min, and then 10°C/min up to 320°C.

MS quality controls for this analysis were performed by injecting an "in-house" standardized plasma pool that was extracted with each batch of 20 samples. Plasma (100 µl) was extracted with methanol and hydroxylamine, and a mixture of internal standards was added. Derivatization was performed using N-methyl-N-tertbutyldimethyl-silyltrifluoroacetamide. Samples were injected onto a GC-MS and operated in electronic ionization mode with helium as reagent gas. Data were corrected for batch effects, and missing values for metabolites were presumed to be below detection range and thus, imputed to be 90% of the minimum detected value. Metabolites were kept if they had ≤ 20% of missing values within each group analyzed.

#### 3. Untargeted Lipidomics

Untargeted lipidomic analysis in the Belgian cohort was conducted in two sequential stages, first a pilot phase using 62 HFpEF and 38 non-HF subjects (2018) and two years later, the main study with 105 HFpEF and 72 non-HF subjects in 2020, with a total of 94 subjects in common between the two phases. Analysis of the Canadian cohort was conducted in 2021 in 74 HFpEF and 103 non-HF subjects. An overview of the study overall workflow for untargeted lipidomics is shown in **Supplementary Figure 1.**

Plasma samples were analyzed using a previously validated label-free semi-quantitative untargeted lipidomic workflow for plasma and serum[41] on a high-resolution LC-MS instruments (LC-quadrupole-time-of-flight (LC-QTOF 6530 and 6550; Agilent Technologies Inc.). MS data were acquired in the positive modes and QC measures were as previously described[6]. In brief, MS data

QCs analyses were performed by (1) injecting an "in-house" plasma pool sample at the beginning, at the end, and every 12 runs; (2) injecting blanks every 20 runs; and (3) monitoring 6 internal standards spiked in samples for signal intensity, mass-to-charge (m/z) ratios, and retention time (RT) accuracies. Raw MS data were processed as previously described in detail using Mass Hunter Qualitative Analysis (version B.06 or B.07; Agilent, Santa Clara, USA) for peak picking and in-house bioinformatic scripts (available upon request on github.com) that were optimized for the following steps using log-2 transformed data: Retention time (RT) correction, normalization of signal intensities using cyclic loess algorithm (with normalizeBetweenArrays function). A maximal value of 20% was set for missing values for a given lipid feature in any groups, and of 80% for the coefficient for inter-individual variation among non-HF subjects. Remaining missing values were imputed using K-nearest neighbors (KNN; with a setting of k=5). Batch correction was performed using Combat algorithm[6].
Raw data from the pilot and main study of the Belgian cohort, as well as from the main study of the Canadian cohort, were processed independently and the resulting final corrected datasets contain 1,637, 1,666 and 1,147 lipid features, respectively, each defined by their m/z, RT and corrected signal intensity. Lipid features were annotated to lipids with their respective acyl chains by MS/MS analysis and with our in-house database as previously described[6]. For lipids represented by multiple features, only the major one was retained. Priority for annotation by MS/MS analysis was given to lipid features found to be significantly associated with HFpEF (see section 3.3). The nomenclature for lipid subclasses generally follows guidelines established by the public database LIPID MAPS. Note that for "oxidized" lipid subclasses, our MS/MS spectra confirmed the presence of an additional oxygen atom on the fatty acyl chain, but did not distinguish between hydroxylated, epoxidized or aldehyde-modified species. This was the case for phosphatidylcholine (PC) species for which the oxidized fatty acyl chain can be hydroxylated or contain an aldehyde group, for triglyceride (TGs) species for which the most common reported oxidation products are the hydroxylated and epoxidized form, and for sphingolipids, for which to our knowledge, only the hydroxylated form has been identified. In the text, these subclasses are abbreviated OxPC, OxTG and OxSL. Corrected datasets between (i) the pilot and main study for the Belgian cohort and (ii) the main study of the Belgian and Canadian cohorts were aligned based on m/z and RT and merged for further data analysis.

After alignment based on mass-to-charge ratio and retention time, 191 annotated lipids were identified as common across both datasets in the overlapping samples (**Pilot-matched,** common across the 94 samples analyzed in both datasets). To assess reproducibility, fold-change estimates for HFpEF versus non-HF subjects were computed independently in the pilot and main studies for each of the 191 shared annotated lipids using identical statistical models. Concordance between studies was evaluated by linear regression of fold-change estimates. This analysis revealed a strong correlation between the two datasets in lipids levels (**Supplementary Figure S2**), demonstrating high reproducibility of lipid effect sizes across independent analytical runs performed two years apart. These results confirm the reproducibility of the untargeted lipidomics workflow over time, and in particular the temporal stability of plasma lipid measurements in frozen samples. This supports the robustness of downstream analyses based on the main study dataset presented in the Results section.

The following two merged corrected datasets were used for subsequent analyses: (1) **Belgian cohort dataset:** 1,666 lipid features with 235 annotated unique lipids. (2) **Cross-cohort dataset:** 158 annotated lipids shared between the Belgian and Canadian cohorts, used for cross-cohort comparisons.

##### 4. Comparison between HF patients and non-HF subjects through unsupervised lipidomic-based analysis

To investigate factors underlying the observed heterogeneity of HFpEF patient global lipid signature, principal component analysis (PCA) was conducted using the 235 annotated unique lipids (**Supplementary Excel File**), which includes the 96 lipids significantly associated with HFpEF (see Results), using R function *prcomp* to reduce dimensions and address collinearity. The PCA representation revealed a PC2-delimited group of 27 HFpEF patients that did not overlap with the 72 non-HF subjects, the subgroup of HFpEF patients with the most distinct lipidomic profiles compared to non-HF (**Figure 1D**. The remaining HFpEF patients (n=78) had lipidomic profiles that were more similar to the non-HF individuals. These two HFpEF subgoups, thereafter referred to as "distinct" (n=27) and "non-distinct" (n=78) HFpEF patients, were compared for their lipid levels with non-HF subjects using pairwise differential analysis. Specifically, three two-way comparisons were made using linear models, with age, sex, and BMI as covariates: (a) non-HF vs. distinct HFpEF. (b) non-HF vs. non-distinct HFpEF. (c) non-distinct HFpEF vs. distinct HFpEF. To account for multiple testing, we computed FDR using *p.adjust* with Benjamini & Hochberg method. The number of lipids significantly (FDR<0.05) associated with distinct HFpEF was found to be greater to that associated to all HFpEF subjects (cf. Results Section 4.1), while non-distinct HFpEF yielded fewer significant results (**Supplementary Figure S3A**) and consistently lower level of significance for the significantly associated lipids (**Supplementary Figure S3B**).

Given that obesity is a recognized risk factor of HFpEF[7] and a major distinctive demographic characteristic of previously documented "obese metabolic" HFpEF phenotype, we further explore the impact of BMI on their lipidomic profile. We used a linear model including an interaction term between categories (non-HF, distinct HFpEF, and non-distinct HFpEF subjects) and BMI, with age and sex as covariates.

Although BMI had little impact on the FC values of the 235 annotated plasma lipids when comparing HFpEF to non-HF subjects (**Supplementary Figure S4A**), we identified with significant interactions effects between BMI and groups (stratified into 3 categories: non-HF, distinct and non-distinct HFpEF) for four lipids (FDR <0.05) namely PC(18:2_22:6), PE(O-16:0/20:4), PC(18:2_22:5), and TG 50:2; **Supplementary Figure S4B),** specifically in the distinct HFpEF subgroup, suggesting that BMI-related lipid signatures are modulated differently among the HFpEF patient population. Altogether, these results highlight the heterogeneity of the lipid signature among HFpEF patients and indicate that a small distinct HFpEF patient group is driving the lipidomic signature of all HFpEF patients, when compared to non-HF subjects, with only a marginal impact of BMI.

#### 5. Consensus clustering analysis

To evaluate the robustness of the HFpEF phenogroups identified by hierarchical clustering in the primary cohort, we performed an independent consensus clustering analysis using the ClustOmics framework[8], an ensemble clustering approach designed to reconcile cluster assignments across multiple algorithms and parameter settings. Importantly, the purpose of the consensus clustering was not to redefine phenogroups, but to assess whether the overall patient stratification identified by hierarchical clustering could be recovered using an algorithm-agnostic strategy. Following batch correction and $\log_2$ transformation of lipid features, multiple clustering solutions were generated using hierarchical clustering (HC), K-means, and partitioning around medoids (PAM), applied both to the full lipidomic space and to a reduced representation obtained by principal component analysis (first 16 components explaining ~80% of the variance). Clusterings were generated for solutions ranging from two to five clusters, yielding a total of 20 input partitions:

- 4 PAM clusterings (2 to 5 clusters) on log2-transformed data
- 4 PAM clusterings (2 to 5 clusters) on reduced data (16 first PC)
- 4 K-means clusterings (2 to 5 clusters) on log2-transformed data
- 4 K-means clusterings (2 to 5 clusters) on reduced data (16 first PC)
- 4 HC clusterings (2 to 5 clusters) on reduced data (16 first PC)

These were integrated using ClustOmics with predefined minimum cluster and consensus sizes to ensure stability. The consensus solution showed strong concordance with the original hierarchical clustering, with a three-cluster structure providing the best balance between modularity and interpretability. Patients overlap between original and consensus clusters was high, and visualization using PCA demonstrated near-identical spatial separation of phenogroups. More than 85% of patients were assigned to the same phenogroup as in the hierarchical clustering, with the highest-risk group showing 24/30 overlap despite small differences in cluster size. Importantly, patients classified into this group by consensus clustering retained the same distinguishing clinical and biochemical features observed in the primary analysis, including markers of cardiometabolic dysfunction and disease severity, and exhibited significantly worse event-free and overall survival when assessed using Kaplan-Meier analysis for mortality from all causes and HF hospitalization (p-value = 0.034). Given this high level of concordance and because hierarchical clustering is more commonly used, we retained the hierarchical clustering solution for all primary analyses and figures. The consensus clustering analysis is presented as a robustness and validation analysis, supporting the stability of the identified HFpEF phenogroups. Of note, this consensus clustering analysis was conducted on an earlier quality-controlled version of the lipidomic dataset, prior to the final refinement of feature filtering used in the main analyses.

#### 6. Comparison of HFpEF clusters between Belgian and Canadian Cohorts

The secondary Canadian cohort includes non-HF subjects (n=103) and HFpEF patients (n=74) from the MHI Biobank with baseline characteristics that differed from the primary Belgian cohort. HFpEF patients in the Canadian cohort had a higher BMI and a greater proportion of males, while non-HF subjects had more CV risk factors (**Supplementary Tables S1** *vs* Table 1 in Pouleur et al. 2024). Similar results were obtained for levels of plasma amino and organic acids (**Supplementary Tables S5)**.

To investigate the overlap in plasma lipidomic data between the two cohorts, we focused on 158 annotated lipids common to both datasets (Dataset 3, **Supplementary Excel File**). Hierarchical clustering was conducted on plasma lipidomic data of HFpEF patients of the Canadian cohorts, which generated also a stable three-cluster solution. There were significant differences amongst the three HFpEF patient clusters of the Canadian cohort for available baseline and biochemical parameters, namely for levels of LDL, HDL and triglycerides; higher values for HDL are seen in cluster C3, but for triglycerides in cluster C1 (**Supplementary Tables S6**), as well as for amino and organic acids, notably with highest values for cysteine, proline and valine in cluster C3, but for glutamate and malate in cluster C1 (**Supplementary Table S7**). In both cases, C2 had intermediate values between C1 and C3. Of note, in contrast to the Belgian cohort, HFpEF patients from the three clusters do not show statistical differences in mean BMI, for which mean values were > 31 for all clusters.

We next compared the plasma lipidomic profiles in the different clusters from the Belgian and Canadian cohorts based using the 158 common annotated lipids. As seen for the 235 annotated lipids (Figure 2A), the HFpEF Belgian clusters show distinct separation along PC1, separating B2 and B3, and PC2, with cluster B1 diverging from B2 and B3 (**Supplementary Figure S6A, left panel**). In contrast, in the Canadian cohort (**Supplementary Figure S6A, right panel**), the HFpEF clusters (C1–C3) separate predominantly along PC1, while no clear distinction is seen along PC2. This suggests a different structure of lipidomic variability in the Canadian cohort, with marginal equivalent to cluster B1 of the Belgian cohort. Altogether these findings suggested that the patient cluster groups in the Canadian cohort cannot easily and directly be mapped to those identified in the Belgian cohort, underscoring the heterogeneity between cohorts.

To investigate which HFpEF patient groups are common or unique between cohorts, we merged the lipidomic datasets, ensuring the removal of batch effects (**Methods**). This allowed for a direct comparison of cluster characteristics as defined independently in both cohorts. Integration of the two datasets enabled visualization of HFpEF patients along with non-HF subjects from both cohorts on a single PCA plot (**Supplementary Figure S6B**). Heatmaps were generated using the pheatmap package (v1.0.12) in R, applying hierarchical clustering with the Ward.D method on the lipids profile directly or on a distance matrix derived from Pearson correlation. This also allows correlation analyses across clusters, with HFpEF patient clusters signatures for the two cohorts showing similarity in their global plasma lipidomic profile (**Figure 4A; upper panel**). This similarity becomes most apparent when computing the lipids' average intensity values per cluster and examining their pairwise correlations (**Figure 4A; lower panel**). Specifically, clusters C1 and C2 showed profiles similar to cluster B3, while cluster C3 aligns most closely with B2, both of which are closest to the non-HF patients from the two cohorts. By contrast, no cluster in the Canadian cohort appeared to match the distinctive lipidomic signature of B1, suggesting this subgroup may be specific to the Belgian cohort

To explore further how HFpEF patients from the Canadian cohort align with the lipidomic clusters identified in the Belgian cohort, we built a Random Forest classifier using the 158 lipids common between cohorts in the Belgian cohort (**Supplementary Figure S7**). The classifier (randomForest_4.7-1.2 package in R) was trained on 70% of the Belgian cohort dataset and their corresponding cluster labels, ensuring balanced cluster representation, and the remaining 30% was used to evaluate model performance using accuracy and confusion matrix metrics. This tree-based model is appropriate for our sample sizes and reported to perform well in metabolomics datasets[9,10]. The trained model was then applied to the Canadian cohort HFpEF patients to predict cluster identities based on the 158 annotated common lipids.

Our classifier demonstrated excellent predictive performance when evaluated on the held-out 30% test set. The model achieved an overall accuracy of 90.91% (95% CI: 75.67%–98.08%), significantly higher than the No Information Rate (NIR = 48.5%, $p = 3.026 \times 10^{-7}$). Sensitivity and specificity values were high across all three classes, with a balanced accuracy (average recall) of 90.05%. These results indicate that the selected lipid features capture the lipidomic structure underlying the HFpEF phenogroups and can reliably reproduce cluster assignments in independent samples. Applying this classifier to the Canadian cohort to predict Belgian cluster identities for each HFpEF patient, we confirmed that there is not a good representation of B1 in the Canadian cohort, with only 6 out of 74 patients predicted as belonging to B1 (**Figure 4B**). This indicates that while some HFpEF clusters clearly have common lipidomic profiles across cohorts, the distinctive plasma lipid pattern of cluster B1 is underrepresented in our secondary cohort. This finding has important implications for understanding cohort-specific variations and the generalizability of lipidomic-based HFpEF phenotyping.

#### 7. Minimal Lipid Signature of HFpEF Cluster with Poor Prognosis

Considering that HFpEF cluster B1 shows the worst event-free survival prognosis (Figure 3) and markers of poor clinical condition (Tables 1-2), we aimed to identify a minimal set of lipids that characterizes patients of this specific subgroup from the other subjects of the Belgian cohort. Using the 235 annotated lipids of the entire Belgian dataset we applied a resampling-based LASSO feature-selection framework with internal cross-validation to handle the high collinearity in lipidomic data. Prior to modelling, low-abundance features were filtered out by removing lipids with a mean intensity below the 1st percentile across samples, leaving 232 lipids for the analysis. Lipid intensities were $\log_2$-transformed and standardized before modelling. To assess feature selection stability, the dataset was repeatedly partitioned into stratified training and held-out subsets (70% training / 30% held-out), ensuring a balanced representation of the B1 class. This procedure was repeated across 200 resampling iterations. In each iteration, all remaining lipids (n=232) were included as predictors in the LASSO-regularized logistic regression model (glmnet, alpha=1) fitted on the training data. The regularization parameter lambda, controlling the strength of the LASSO penalty, was selected by internal cross-validation within the training set, and predictors with non-zero coefficients were recorded at each iteration. Lipids selected in more than 50% of the 200 training iterations were retained as the candidate minimal lipid signature associated with the B1 cluster. This resampling-based stability selection was designed to prioritize lipid features that are consistently selected across data partitions rather than to derive a finalized predictive classifier. We then fitted a ridge logistic regression model using these lipids to generate a continuous B1-score (fitted probability) used for visualization (**Figure S8A**).

This approach identified a set of 10 lipids from 6 different subclasses, for which the PCA projection showed a clear and distinct positioning of B1 in the lipidomic signature space vs. other patient clusters, thereby supporting their discriminative power (**Figure S8A**) with patients colored by their probability of belonging to cluster B1 based on the ridge regression model. Given the strong correlation structure observed in plasma lipidomics, these lipids should be interpreted as representatives of broader correlated lipid modules rather than as uniquely causal biomarkers. These 10 lipids were correlated with each other either positively or negatively (**Figure 5A**) and also showed a strong correlation network with all other lipids, including 8 additional lipids selected in more than 30% of iterations (**Figure S8B**), with an overall pattern closely resembling that of **Figure 1C**. Heatmap visualization of the retained lipids was performed using the *pheatmap* package (v1.0.12) in R with hierarchical clustering based on the Ward.D linkage method. We performed a network analysis based on pairwise correlations anchored in all lipids used in the minimal signature identification step, and visualized correlations (r>0.6) using the Fruchterman-Reingold algorithm using the package *igraph* in R (v2.0.3).

Differences of signal intensities between clusters for these 10 lipids, shown as box plots (**Figure 5B**), highlight that B1 patients not only differ from clusters B2 and B3, but also from non-HF subjects, with their plasma levels being either higher (CAR 18:1, CAR 26:1, 1 oxSM, 1 ether PC) or lower (3 PCs, 1 ether PE, 1 TG, all with PUFA acyl chains, as well as 1 SM with a VLC-FA), reinforcing the distinctiveness of this subgroup. Finally, we tested the relationship between these 10 lipids with clinical and biochemical parameters among all HFpEF patients (**Figure 5C; Figure S9**) using two complementary correlation approaches based on variable type: Spearman's rank correlation was applied to continuous clinical variables, and point-biserial correlation was used for binary variables. This mixed-method approach ensures appropriate handling of both data types. Notably, most of the parameters are associated with markers of disease severity, namely reflecting cardiac and liver necrosis or fibrosis (TnT, FGF-23, ST2, Fib-4, NAFLD score), congestion (CA-125), either positively (CAR 18:1 and 26:1, OxSM, PC(O-20:0/20:4)) or negatively (3PC with a PUFA, PE(O-16:0/22:6), SM(d18:1/21:0)), which are all distinctive clinical characteristics of cluster B1. Thus, this analysis provided additional support for the notion that these 10 lipids are differentially associated with specific clinical and biochemical parameters that discriminate HFpEF patient cluster B1 from other clusters.

#### 8. Supplementary Text References

**Supplementary Excel Files with lipidomic data**

**1) Supplementary Excel File 1 – Includes 4 data sheets related to Figures.**

- **Data Sheet 1 – related to Figure 1A-C & S2**: Comparison of intensity signal values for plasma lipid features between HFpEF patients vs. non-HF subjects in the Belgian cohort: Results for the independent testing on the 1,666 lipid features in HFpEF patients vs. non-HF subjects.

- **Data Sheet 2 – Related to Figure 1D, 2, 4-5, S3-S7:** Corrected intensity signal values of the 235 annotated unique lipids for HFpEF patients according to clusters and non-HF subjects in Belgian cohort.

- **Data Sheet 3 – Related to Figure 2C & S5:** Statistical results for the comparison of the 235 annotated unique lipids between HFpEF patients from different clusters in Belgian cohort using Kruskall Wallis test.

- **Data Sheet 4 – Related to Figure 4-5 & S6-S7:** Corrected intensity signal values for the 158 annotated unique lipids common to the Belgian and Canadian datasets for HFpEF patients according to different clusters and non-HF patients in the Canadian cohort).

**2) Mendeley dataset – Includes 3 sheets with corrected datasets for the Belgian cohort pilot (Sheet 1) and main (Sheet 2) studies and for Canadian cohort (Sheet 3).**

#### Supplementary Figures

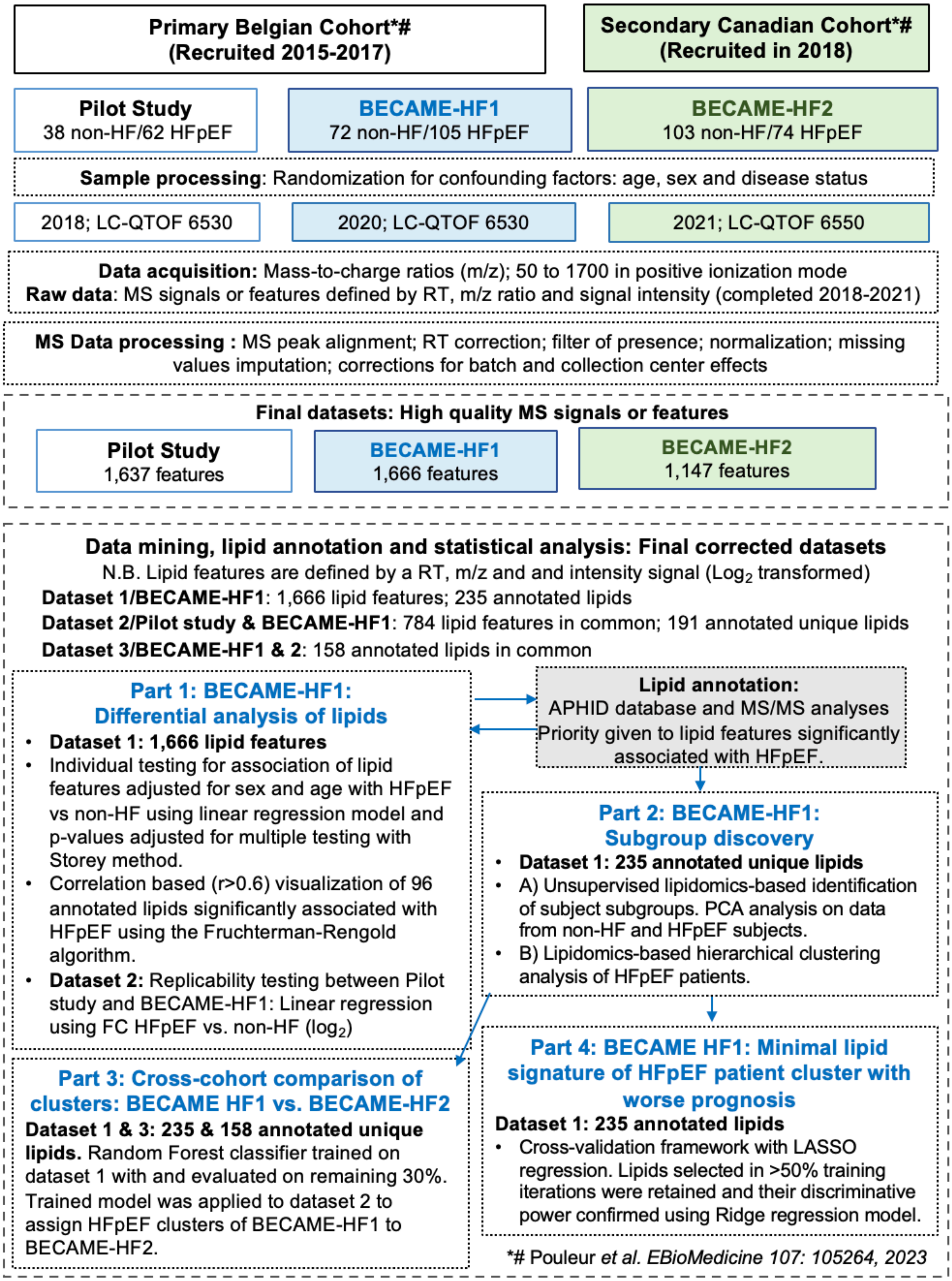


**Figure S1: Workflow diagram for untargeted lipidomic analysis in BECAME-HF project.**

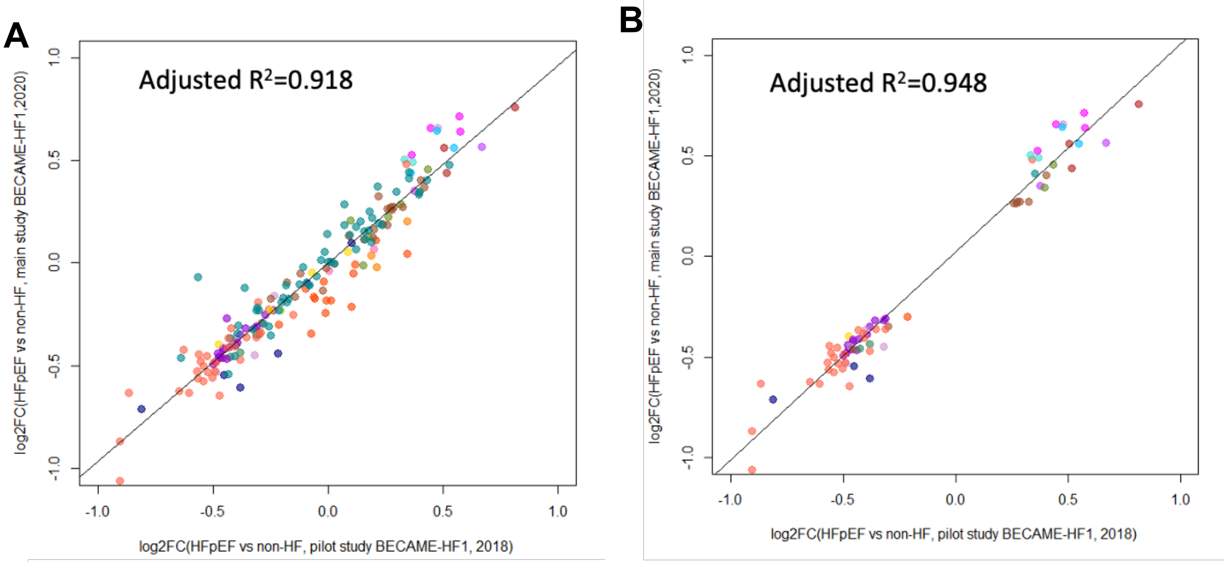


**Figure S2. Reproducibility of lipidomic results between pilot and main studies in the Belgian cohort.** Comparison of fold-change estimates obtained in the pilot and main studies using the 94 samples analyzed in both datasets. Data are expressed as $log_2$ fold-change in lipid intensity for HFpEF versus non-HF subjects in the pilot study (x-axis) versus the main study (y-axis). **(A)** All 191 annotated lipids detected in both datasets. **(B)** Subset of 75 annotated lipids significantly associated with HFpEF in the main study ($p < 0.01$). Linear regression demonstrates strong concordance of effect sizes across studies (adjusted $R^2$ shown). Among these shared lipids, 72, 61, and 89 were significantly different when comparing HFpEF clusters B1, B2, and B3, respectively, with the other two clusters.

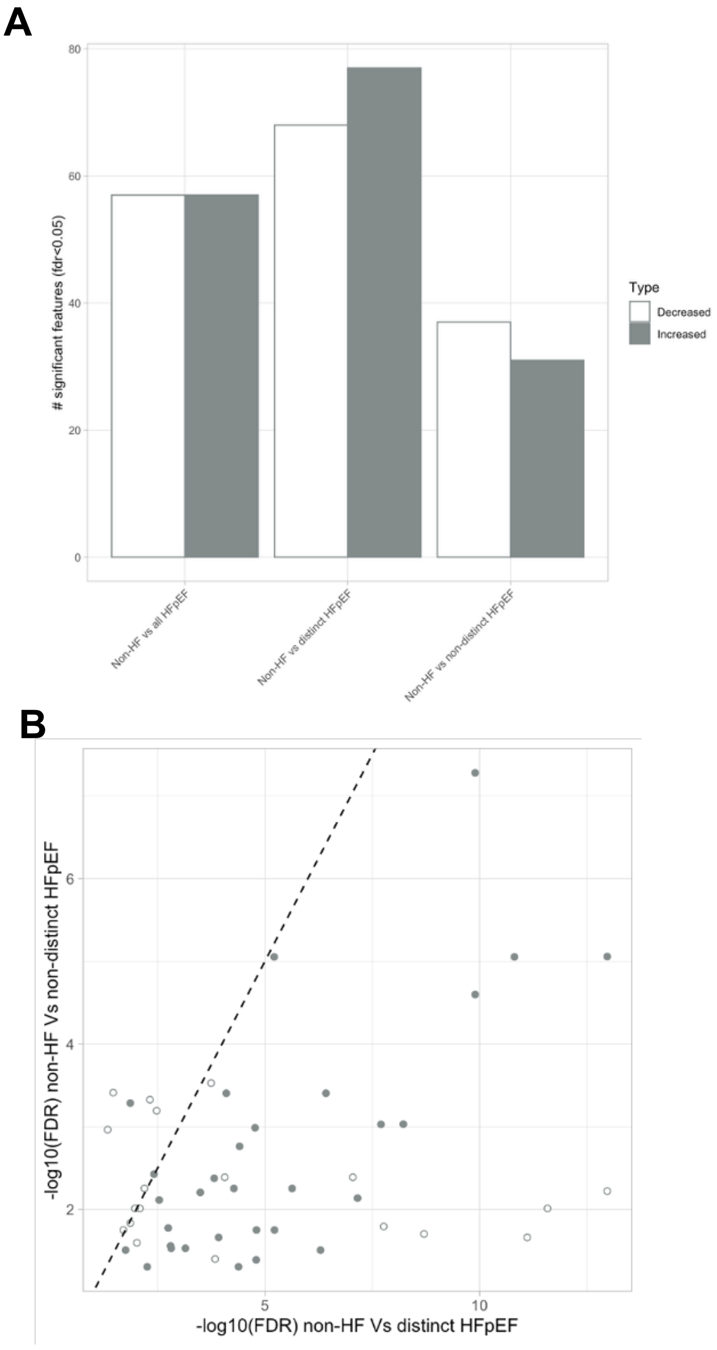


**Figure S3: Comparisons using pairwise differential analysis on the 235 annotated plasma lipids between all HFpEF, distinct and non-distinct HFpEF group vs. non-HF subjects in Belgian cohort. (A)** Bar graphs showing the number of significant unique lipids for the various comparisons. **(B)** Linear relationship between FDR for (i) non-HF vs. distinct HFpEF and non-HF vs. "non-distinct" HFpF.

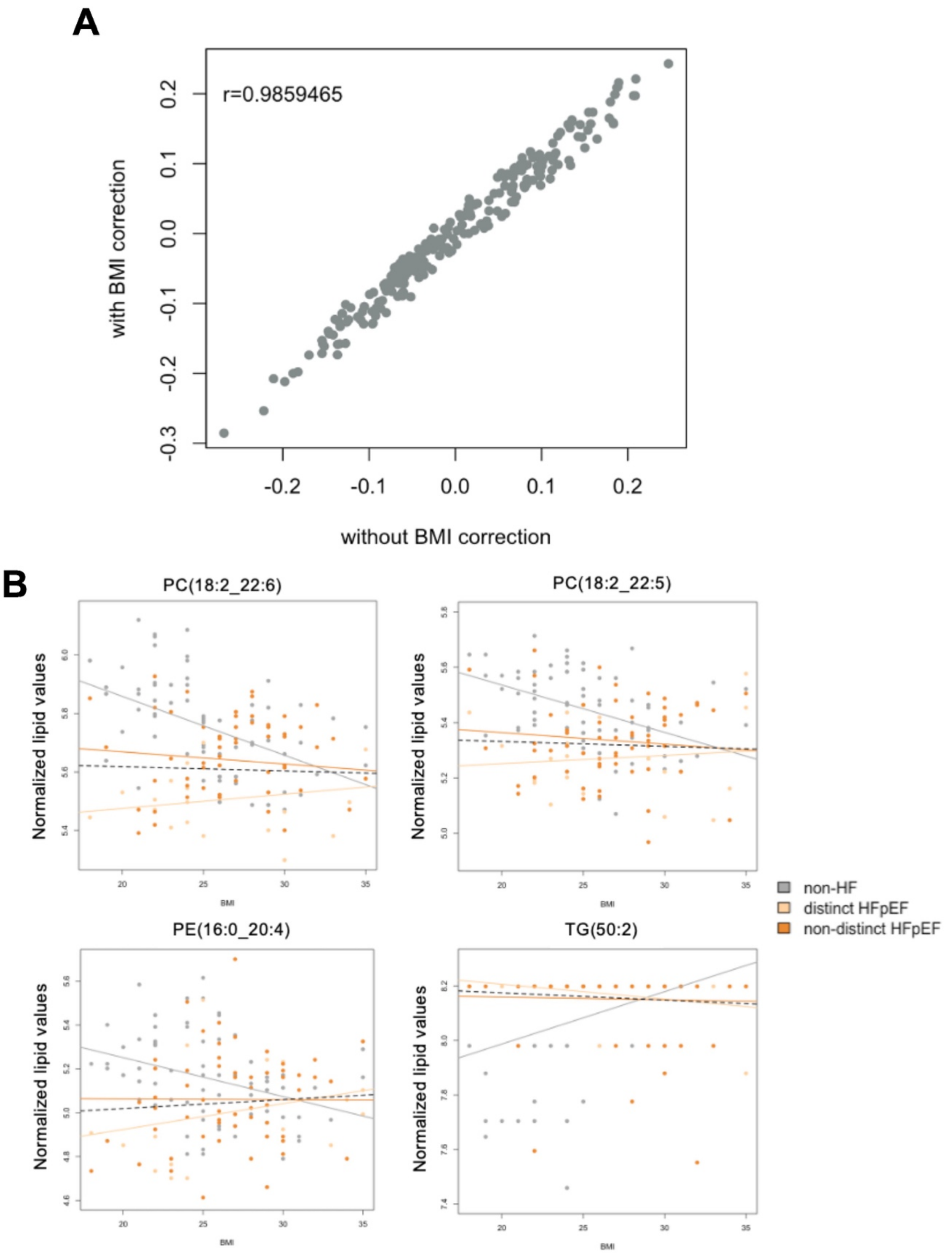


**Figure S4: Impact of BMI on the 235 annotated lipids in Belgian cohort. (A)** Intensity values of lipids of HFpEF vs. non-HF expressed in fold-change (log2) with or without BMI correction. **(B)** Intensity values of lipids showing a significant interaction between BMI and subject groups (non-HF, non-distinct and distinct HFpEF)

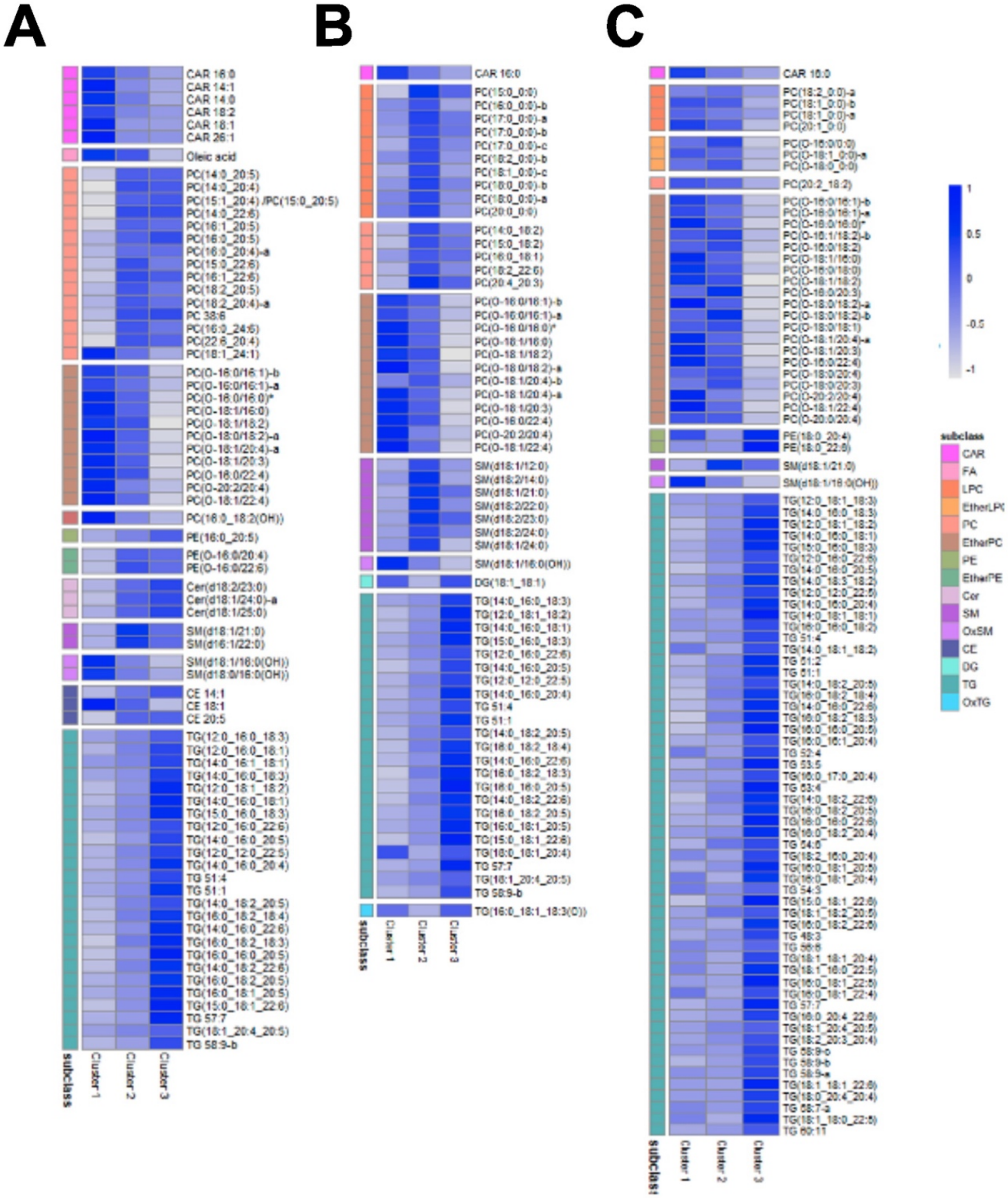


**Figure S5**: **Comparison of plasma lipids between HFpEF patient clusters from the Belgian cohort shown as a heatmap. (A)** Cluster 1 vs. 2 and 3; **(B)** Cluster 2 vs. 1 and 3; **(C)** Cluster 3 vs. 1 and 2. (Criteria for inclusion: Kruskal Wallis p-value $< 7 \times 10^{-4}$ and Wilcoxon pair-wise comparison p-value<0.02)

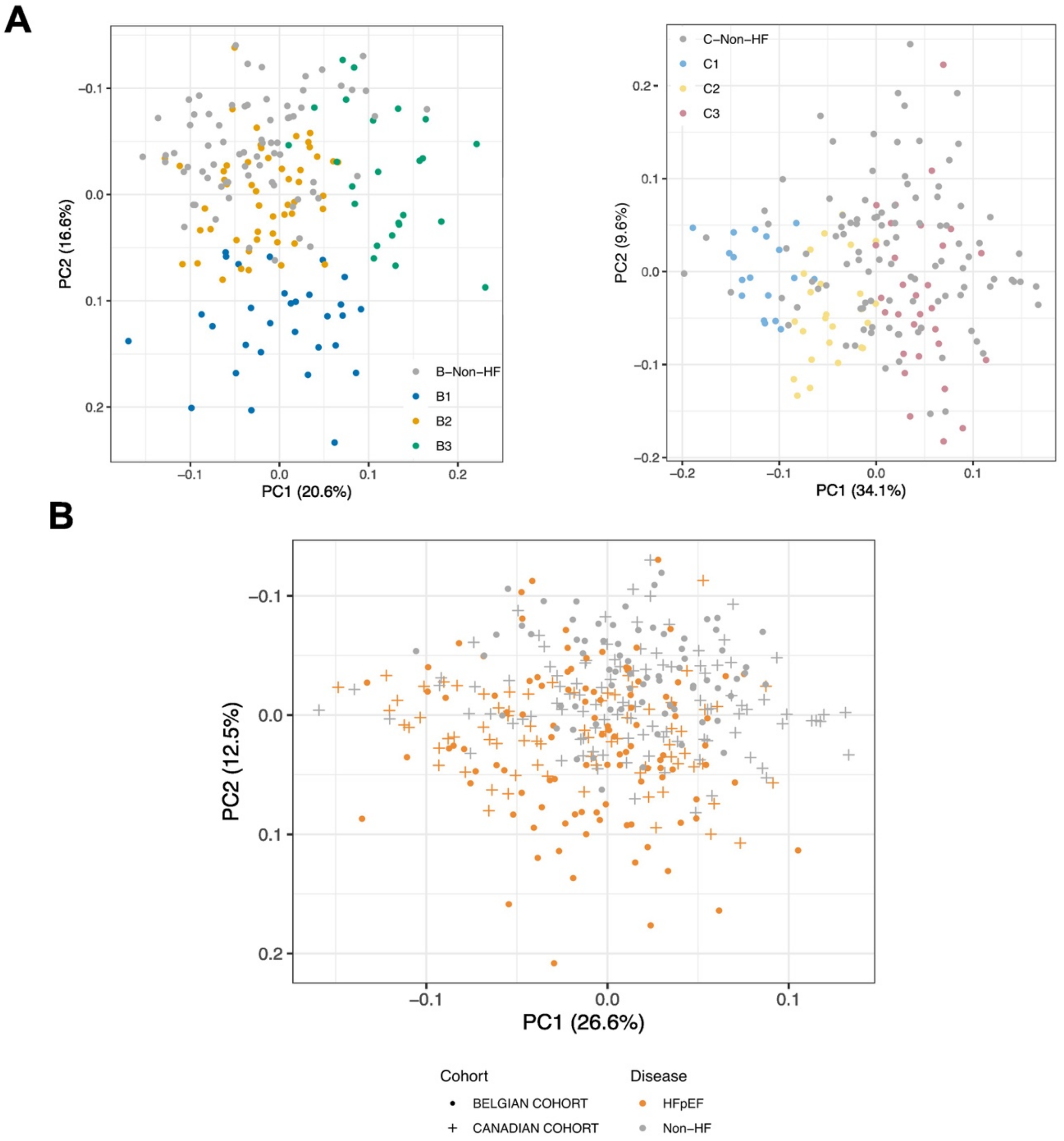


**Figure S6**: **Visualization of datasets containing 158 common annotated unique lipids between Belgian (B) and Canadian (C) cohorts (A) before and (B) after integration into a share space. (A)** PCA score plot of intensity value data after log transformation and scaling for the 158 common annotated plasma lipids of non-HF subjects and HFpEF patients with inferred clusters for the Belgian (B; left panel) and Canadian (C; right panel) cohorts; **(B)** PCA score plot showing the integrated dataset from the Belgian and Canadian cohorts after concatenation, $log_2$-transformation and batch effect correction.

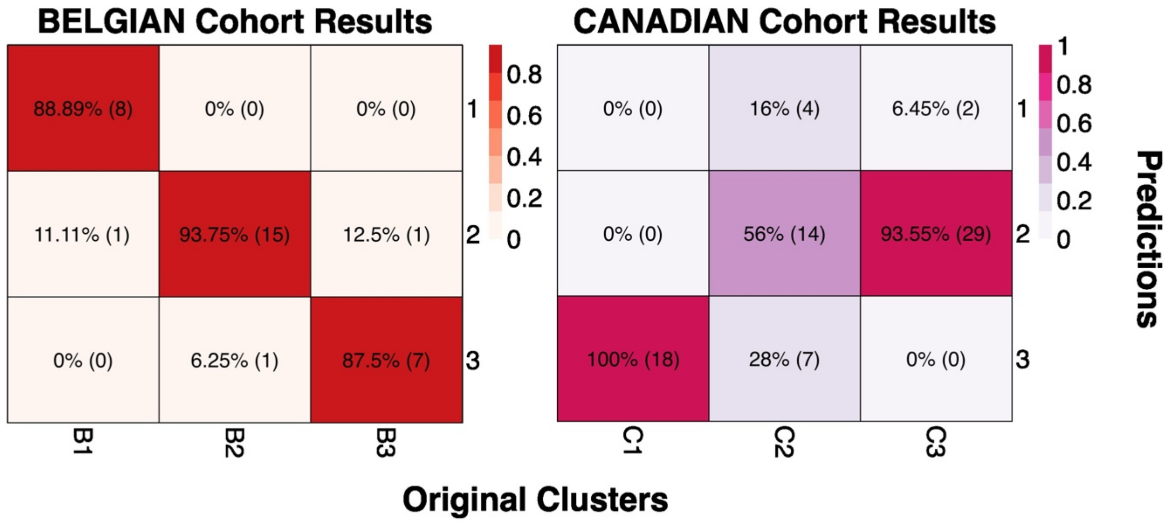


**Figure S7: Performance of classification model using Random Forest for HFpEF cluster prediction in Belgian and Canadian cohorts.**

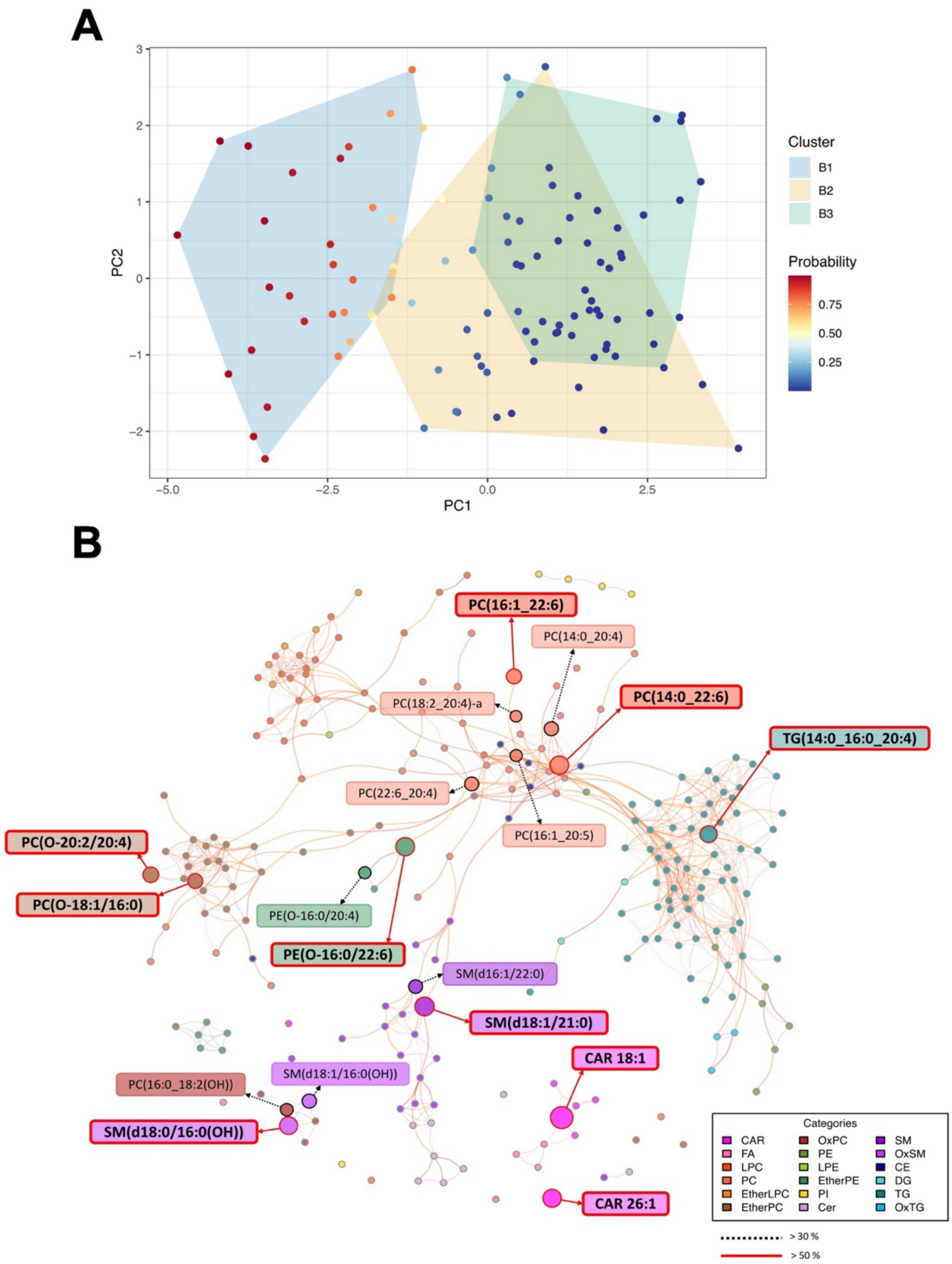


**Figure S8: Identification using a cross-validation approach with Lasso regression of a minimal signature for distinguishing HFpEF cluster with worse survival prognosis in the Belgian cohort. (A)** PCA score plot colored by probability of prediction using the ridge regression model. **(B)** Correlation network for the annotated BECAME-HF1 lipids, highlighting those with 50% and 30% frequency. Node sizes: Frequency in CV; Node colors: Lipid subclass; Labels: Frequency <= 30% (grey border), Frequency >30% (black border), Frequency >50% (red border)

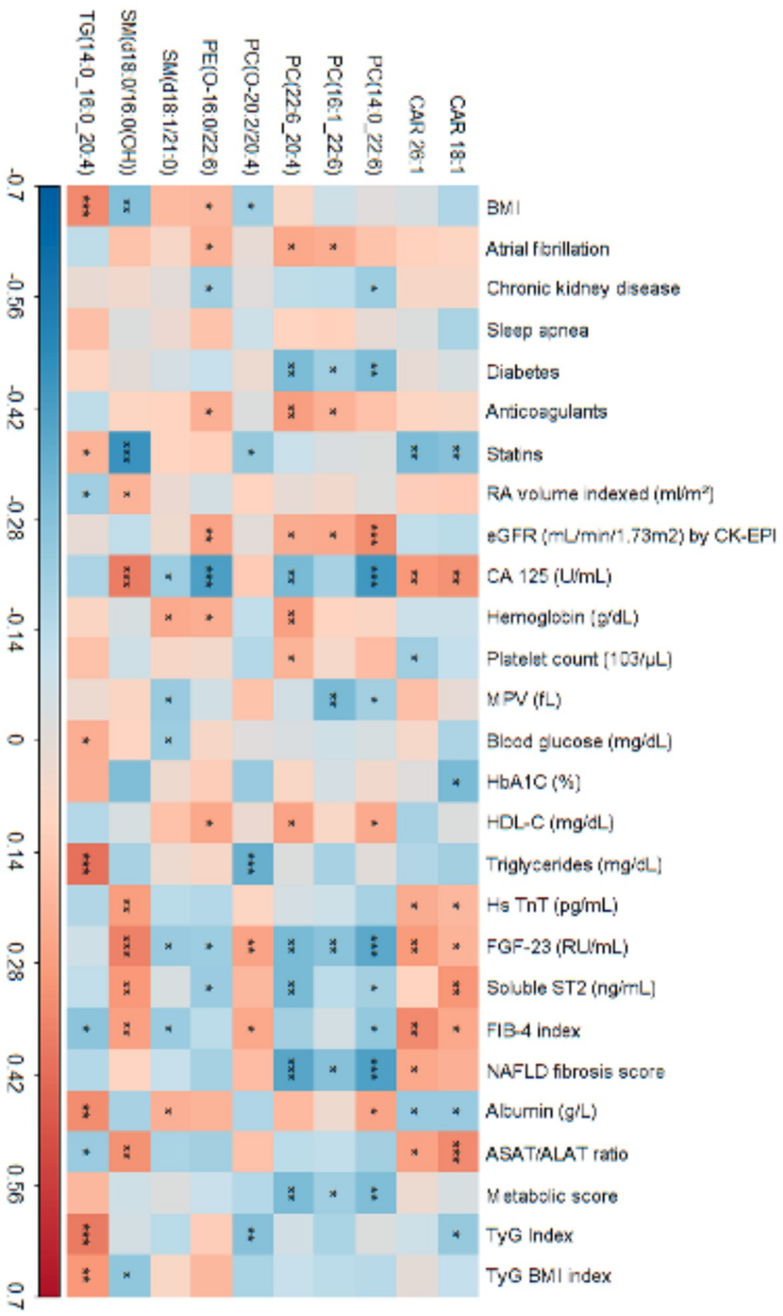


**Figure S9: Heat map illustrating the correlation between lipids of the minimal signature with clinical and biological parameters of HFpEF patients in Belgian cohort.** Spearman correlation for continuous variables; point-biserial for categorical variables.

#### Supplementary Tables

**Supplementary Table S1- Related to Figure 1.** Baseline characteristics of non-HF and HFpEF subjects in Belgian cohort.

**Supplementary Table S2 - Related to Figure 1.** Plasma levels of organic and amino acids in non-HF and HFpEF subjects in Belgian cohort.

**Supplementary Table S3 - Related to Table 2.** Targeted analysis of plasma amino and organic acids in HFpEF patients of the Belgian cohort according to cluster subgroup.

**Supplementary Table S4 - Related to Figure 3 and Table 1**. Cox regression of the secondary endpoint (mortality from all-causes) for HFpEF patient clusters**.**

**Supplementary Table S5 – Related to Supplemental Table S2.** Plasma levels of organic and amino acids in non-HF and HFpEF subjects in Canadian cohort.

**Supplementary Table S6 – Related to Table 1 and 2.** Baseline characteristics in HFpEF patients of the Canadian cohort according to cluster subgroups.

**Supplementary Table S7 – Related to Supplemental Table S3.** Targeted analysis of plasma amino and organic acids in HFpEF patients of the Canadian according to cluster subgroups.

**Supplementary Table S1.** Baseline characteristics of Belgian cohort non-HF and HFpEF subjects.

| | **Non-HF** | **HFpEF** | ***P value*** |
|---|---|---|---|
| **Baseline characteristics** | N = 72 | N = 105 | |
| **Age (years)** | 63±12 | 76±7 | <0.001 |
| **Female (*n*, %)** | 33 (46) | 60 (57) | 0.185 |
| **BMI (kg/m$^2$)** | 26±4 | 29±7 | <0.001 |
| **Heart rate (beat/min)** | 67±10 | 71±14 | 0.027 |
| **Systolic blood pressure (mmHg)** | 140±19 | 137±22 | 0.461 |
| **Diastolic blood pressure (mmHg)** | 82±11 | 74±14 | <0.001 |
| **NYHA class III and IV (*n*, %)** | 0 (0) | 46 (44) | <0.001 |
| **Medical history** | | | |
| **Atrial fibrillation (*n*, %)** | 0 (0) | 68 (65) | <0.001 |
| **Ischemic cardiomyopathy (*n*, %)** | 0 (0) | 41 (39) | <0.001 |
| **COPD (*n*, %)** | 0 (0) | 12 (11) | 0.012 |
| **Sleep apnea (*n*, %)** | 0 (0) | 16 (15) | 0.003 |
| **Cardiovascular risk factors** | | | |
| **Hypertension (*n*, %)** | 27 (38) | 97 (92) | <0.001 |
| **Diabetes (*n*, %)** | 14 (19) | 47 (45) | 0.001 |
| **Hypercholesterolemia (*n*, %)** | 49 (68) | 68 (65) | 0.836 |
| **Smoking (*n*, %)** | 16 (22) | 49 (47) | <0.001 |
| **Family history of CV disease (*n*, %)** | 13 (18) | 22 (21) | 0.753 |
| **Medication** | | | |
| **ACE inhibitor - ARBs (*n*, %)** | 18 (25) | 72 (69) | <0.001 |
| **Beta blocker (*n*, %)** | 4 (6) | 74 (71) | <0.001 |
| **Loop diuretics (*n*, %)** | 0 (0) | 74 (71) | <0.001 |
| **Thiazide (*n, %)*** | 4 (6) | 23 (22) | 0.006 |
| **MRA (*n*, %)** | 0 (0) | 20 (19) | <0.001 |
| **Anticoagulants (*n*, %)** | 0 (0) | 60 (57) | <0.001 |
| **Antiplatelet agents (*n*, %)** | 13 (18) | 46 (44) | <0.001 |
| **Statins (*n*, %)** | 16 (22) | 50 (49) | <0.001 |
| **Echocardiography study** | | | |
| **LA volume indexed (ml/m$^2$)** | 21±6 | 45±18 | <0.001 |
| **LV ejection Fraction (%)** | 64±5 | 62±8 | 0.028 |
| **E/e' septal ratio** | 10±3 | 19±8 | <0.001 |
| **eSPAP (mmHg)** | 18±5 | 32±10 | <0.001 |
| **Biology** | | | |
| **NT-proBNP (pg/mL)** | 57 [42;116] | 1927 [988;3442] | <0.001 |
| **eGFR (mL/min/1.73m$^2$) by CK-EPI** | 74±16 | 55±20 | <0.001 |
| **Hemoglobin (g/dL)** | 14 ±1 | 12±2. | <0.001 |
| **Neutrophil-to-lymphocyte ratio** | 2.0 [1.7;2.3] | 3.3 [2.4;5.0] | <0.001 |
| **Platelet count (10$^3$/µL)** | 255 [196;283] | 224 [185;286] | 0.208 |
| **MPV (fL)** | 9.9 [9.5;10.3] | 10.6 [9.8;11.3] | 0.001 |
| **Total cholesterol (mg/dL)** | 195 [166;220] | 140 [117;169] | <0.001 |
| **LDL-C (mg/dL)** | 111 [91;136] | 69 [50 ;94] | <0.001 |
| **HDL-C (mg/dL)** | 62 [53;73] | 50 [40;59] | <0.001 |
| **Triglycerides (mg/dL)** | 96 [66;118] | 92 [72;120] | 0.846 |
| **CRP (mg/dL)** | 0.20 [0.10;0.30] | 0.75 [0.23;2.38] | <0.001 |
| **Hs TnT (pg/mL)** | 6.0 [4.0;8.0] | 28.0 [16.0;35.0] | <0.001 |
| **FGF-23 (RU/mL)** | 61 [47;77] | 243 [122;546] | <0.001 |
| **Soluble ST2 (ng/mL)** | 26 [22;32] | 42 [31;62] | <0.001 |
| **Albumin (g/L)** | 45 [43;47] | 39 [36;42] | <0.001 |

ACE, angiotensin-converting enzyme; ARB, angiotensin receptor blocker; BMI, body mass index; COPD, chronic obstructive pulmonary disease; CRP, C-reactive protein; CV, cardiovascular; eGFR, estimated glomerular filtration rate; eSPAP, estimated systolic pulmonary artery pressures; FGF-23, fibroblast growth factor 23; HF, heart failure; HFpEF, heart failure with preserved ejection fraction; HFrEF, heart failure with reduced ejection fraction; HDL-C, high-density lipoprotein cholesterol; hsTnT, high-sensitivity troponinT; LA, left atrium; LDL-C, low-density lipoprotein cholesterol; LV, left ventricular; MRA, mineralocorticoid receptor antagonist; MPV, mean platelet volume; NT-proBNP, N-terminal pro B-type natriuretic peptide; NYHA, New York Heart Association; SBP, systolic blood pressure; ST2; suppression of tumorigenicity 2.

Values are mean ± standard deviation or median and interquartile range (IQR 0.25-0.75). Categorical variables are expressed as count and proportion. Differences between clinical characteristics were compared using independent T-test or chi-squared test where appropriate.

**Supplementary Table S2.** Targeted analysis of plasma amino and organic acids in a subset of non-HF and HFpEF subjects from the Belgian cohort.

| | **Non-HF** | **HFpEF** | **Fold-change** | ***P value*** |
|---|---|---|---|---|
| | N= 44 | N=70 | | |
| **Amino acids (µM)** | | | | |
| Alanine | 372.4 ± 122.7 | 367.1 ± 109.4 | 0.96 | 0.580 |
| Arginine | 189.0 ± 61.6 | 198.7 ± 72.8 | 1.02 | 0.738 |
| primaryAsparagine | 37.5 ± 9.7 | 38.2 ± 10.0 | 0.98 | 0.803 |
| Aspartate | 5.3 ± 2.5 | 5.1 ± 2.7 | 0.95 | 0.626 |
| **Cysteine** | **158.2 ± 38.7** | **221.1 ± 79.5** | **1.32** | $\mathbf{1.04 \times 10^{-6}}$ |
| Glutamate | 102.0 ± 73.7 | 107.4 ± 88.5 | 1.05 | 0.720 |
| Glutamine | 513.5 ± 98.9 | 529.5 ± 123.8 | 0.98 | 0.918 |
| Glycine | 214.8 ± 53.6 | 226.3 ± 61.8 | 1.05 | 0.347 |
| Histidine | 75.8 ± 8.0 | 70.3 ± 11.7 | 0.92 | 0.025 |
| **Hydroxyproline** | **2.0 ± 0.5** | **2.7 ± 1.2** | **1.33** | $\mathbf{8.19 \times 10^{-5}}$ |
| Isoleucine | 76.1 ± 22.8 | 84.2 ± 33.5 | 1.10 | 0.163 |
| Leucine | 106.6 ± 25.5 | 114.0 ± 39.3 | 1.05 | 0.416 |
| Lysine | 181.2 ± 38.4 | 200.8 ± 58.6 | 1.08 | 0.199 |
| Methionine | 22.2 ± 5.7 | 24.7 ± 8.7 | 1.01 | 0.823 |
| Phenylalanine | 60.6 ± 9.4 | 68.3 ± 14.0 | 1.07 | 0.085 |
| Proline | 213.8 ± 80.4 | 237.1 ± 76.8 | 1.17 | 0.026 |
| Serine | 87.4 ± 18.2 | 84.2 ± 20.8 | 0.96 | 0.437 |
| Threonine | 124.7 ± 27.5 | 124.8 ± 40.9 | 0.96 | 0.545 |
| Tryptophan | 59.5 ± 16.3 | 56.3 ± 20.6 | 0.90 | 0.150 |
| Tyrosine | 68.5 ± 14.8 | 73.7± 24.7 | 0.98 | 0.780 |
| Valine | 241.4 ± 44.8 | 251.3 ± 66.4 | 1.04 | 0.429 |
| **Organic acids (µM)** | | | | |
| Acetoacetate | 10.8 ± 15.6 | 12.3 ± 19.8 | 1.33 | 0.422 |
| α-Hydroxybutyrate | 1415.8 ± 607.7 | 1456.3 ± 740.1 | 0.92 | 0.360 |
| α-Ketobutyrate | 4.2 ± 2.3 | 4.6 ± 2.9 | 0.95 | 0.767 |
| β-Hydroxybutyrate | 112.5 ± 126.6 | 86.8 ± 100.2 | 0.67 | 0.074 |
| Citrate | 103.7 ± 20.6 | 105.6 ± 30.8 | 0.92 | 0.205 |
| α-Ketoglutarate | 26.6 ± 8.8 | 34.2 ± 14.1 | 1.30 | 0.056 |
| Lactate | 1517.9 ± 720.9 | 1788.5 ± 854.7 | 1.21 | 0.055 |
| Malate | 8.9 ± 4.2 | 9.6 ± 2.8 | 1.07 | 0.383 |
| Pyruvate | 216.1 ± 138.9 | 276.5 ± 135.3 | 1.46 | 0.029 |
| Succinate | 17.8 ± 6.6 | 18.3 ± 3.7 | 1.01 | 0.907 |
| **Calculated ratios** | | | | |
| Lactate/Pyruvate | 10.7 ± 9.9 | 9.2 ± 10.8 | 0.83 | 0.243 |
| β-Hydroxybutyrate/Acetoacetate | 130.3 ± 321.24 | 54.6 ± 98.4 | 0.51 | 0.063 |
| α-Hydroxybutyrate/α-Ketobutyrate | 426.9 ± 336.09 | 433.5 ± 443.5 | 0.96 | 0.772 |
| Glutamine/Glutamate | 9.1 ± 7.22 | 8.1 ± 5.4 | 0.93 | 0.803 |

Values are mean ± standard deviation. Metabolites shown in this table had ≤ 20% missing values within each group (i.e. HFpEF or controls). Missing values were imputed as 90% of the minimal value of each metabolite. Fold-changes and *P*-values were calculated using linear regression with batch, age and sex as covariates. Metabolite shown in bold are statistical significant for HFpEF vs. Non-HF using as threshold ≤0.01 and absolute fold-change │FC │>1.25.

**Supplementary Table S3.** Targeted analysis of plasma amino and organic acids in HFpEF patients of the Belgian primary cohort according to cluster subgroup.

| | Cluster 1 | Cluster 2 | Cluster 3 | *P (Levene)* | *P (Anova)* | *P (Kruskal-Wallis)* |
|---|---|---|---|---|---|---|
| | N=17 | N=35 | N=18 | | | |
| **Amino acids (µM)** | | | | | | |
| Alanine | 355.1 ± 95.3 | 378.1 ± 106.4 | 356.9 ± 130.0 | 0.188 | 0.512 | 0.956 |
| Arginine | 194.8 ± 71.3 | 200.0 ± 82.6 | 199.8 ± 55.3 | 0.369 | 0.858 | 0.939 |
| Asparagine | 41.2 ± 10.3† | 39.2 ± 9.3 | 33.3 ± 9.7 | 0.316 | 0.026 | 0.090 |
| Aspartate | 5.7 ± 3.5 | 4.8 ± 2.1 | 5.1 ± 2.9 | 0.527 | 0.724 | 0.878 |
| Cysteine | 217.6 ± 76.2 | 219.4 ± 69.4 | 227.6 ± 102.5 | 0.877 | 0.983 | 0.926 |
| Glutamate | 118.5 ± 107.2 | 106.8 ± 89.8 | 98.3 ± 67.8 | 0.329 | 0.832 | 0.858 |
| Glutamine | 521.4 ± 116.2 | 542.3 ± 129.7 | 512.2 ± 122.9 | 0.964 | 0.844 | 0.349 |
| Glycine | 236.1 ± 69.7 | 227.3 ± 59.5 | 215.1 ± 60.0 | 0.535 | 0.595 | 0.719 |
| Histidine | 69.4 ± 8.8 | 73.5 ± 11.8 | 65.0 ± 12.4 | 0.724 | 0.029 | 0.101 |
| Hydroxyproline | 2.9 ± 1.8 | 2.7 ± 0.9 | 2.6 ± 1.1 | 0.857 | 0.830 | 0.980 |
| Isoleucine | 87.7 ± 32.8 | 83.4 ± 35.1 | 82.5 ± 32.5 | 0.955 | 0.813 | 0.816 |
| Leucine | 119.6 ± 38.5 | 112.0 ± 42.7 | 112.8 ± 34.5 | 0.756 | 0.724 | 0.858 |
| Lysine | 207.9 ± 57.0 | 203.4 ± 61.4 | 189.1 ± 55.9 | 0.546 | 0.502 | 0.640 |
| Methionine | 27.5 ± 11.0 | 24.2 ± 7.9 | 22.9 ± 7.3 | 0.370 | 0.303 | 0.445 |
| Phenylalanine | 70.6 ± 15.8 | 67.7 ± 13.7 | 67.5 ± 13.4 | 0.610 | 0.812 | 0.738 |
| Proline | 238.2 ± 64.7 | 229.3 ± 68.2 | 251.2 ± 101.7 | 0.135 | 0.803 | 0.856 |
| **Serine** | **96.5 ± 27.2†** | **84.6 ± 15.9†** | **71.8 ± 15.6** | **0.160** | **0.004** | **0.003** |
| Threonine | 138.6 ± 43.3† | 127.4 ± 40.6 | 106.7 ± 34.1 | 0.935 | 0.048 | 0.069 |
| Tryptophan | 56.8 ± 22.5 | 59.2 ± 21.3 | 50.2 ± 16.7 | 0.470 | 0.346 | 0.525 |
| Tyrosine | 79.4 ± 28.5 | 73.6 ± 25.3 | 68.4 ± 19.5 | 0.876 | 0.514 | 0.612 |
| Valine | 251.5 ± 68.3 | 248.7 ± 70.1 | 256.5 ± 60.2 | 0.592 | 0.876 | 0.808 |
| **Organic acids (µM)** | | | | | | |
| Acetoacetate | 9.4 ± 11.3 | 10.5 ± 16.5 | 18.5 ± 29.5 | 0.885 | 0.718 | 0.779 |
| α-Hydroxybutyrate | 1365.8 ± 634.2 | 1296.6 ± 585.5 | 1852.2 ± 965.5 | 0.504 | 0.087 | 0.089 |
| α-Ketobutyrate | 4.1 ± 3.1 | 3.9 ± 1.4 | 6.4 ± 4.0 | 0.001 | 0.023 | 0.084 |
| β-Hydroxybutyrate | 81.1 ± 101.4 | 86.7 ± 102.5 | 92.3 ± 100.0 | 0.753 | 0.851 | 0.830 |
| Citrate | 96.2 ± 27.9# | 109.5 ± 27.9 | 107.1 ± 37.8 | 0.195 | 0.405 | 0.154 |
| α-Ketoglutarate | 30.2 ± 11.9 | 35.9 ± 15.0 | 34.9 ± 14.1 | 0.917 | 0.398 | 0.585 |
| Lactate | 1876.7 ± 883.4 | 1813.5 ± 914.0 | 1656.5 ± 729.5 | 0.862 | 0.646 | 0.628 |
| Malate | 10.0 ± 2.2 | 9.9 ± 3.3 | 8.9 ± 1.9 | 0.729 | 0.371 | 0.295 |
| Pyruvate | 244.3 ± 118.5 | 293.2 ± 142.0 | 274.4 ± 138.5 | 0.735 | 0.357 | 0.577 |
| Succinate | 19.0 ± 5.4 | 18.5 ± 2.7 | 17.3 ± 3.5 | 0.031 | 0.378 | 0.435 |
| **Calculated ratios** | | | | | | |
| Lactate/Pyruvate | 15.2 ± 19.7 | 7.2 ± 4.3 | 7.4 ± 4.8 | 0.105 | 0.226 | 0.737 |
| β-Hydroxybutyrate/Acetoacetate | 42.2 ± 84.1 | 62.9 ± 110.7 | 50.1 ± 88.3 | 0.659 | 0.848 | 0.894 |
| α-Hydroxybutyrate/α-Ketobutyrate | 687.5 ± 824.5 | 360.5 ± 166.4 | 335.3 ± 132.3 | 0.073 | 0.126 | 0.680 |
| Glutamine/Glutamate | 7.8 ± 5.4 | 7.9 ± 4.7 | 9.0 ± 6.8 | 0.591 | 0.945 | 0.928 |

Values are mean ± standard deviation. Missing values were imputed as 90% of the minimal value of each metabolite. Statistic tests were performed on log2-transformed data. Equality of variance between clusters was tested by Levene test. Metabolite shown in bold is statistically significant with $P \leq 0.05$ following Kruskal-Wallis test.

### $p<0.05$ individual category vs. cluster 2 following Wilcoxon test

† $p<0.05$ individual category vs. cluster 3 following Wilcoxon test

**Supplementary Table S4**. Cox regression for the secondary endpoint (mortality from all-causes) for HFpEF patient clusters in Belgian cohort

| **Cox regression analysis**<br>**Primary endpoint** | **Univariable** | | **Multivariable** | |
|---|---|---|---|---|
| | Hazards ratio 95% CI | *P-value* | Hazards ratio 95% CI | *P-value* |
| Cluster 1 vs 2 and 3 | 2.20 (1.37-3.54) | 0.001 | 1.98 (1.19-3.82) | 0.008 |
| Cluster 2 vs 1* | 0.46 (0.28-0.77) | 0.003 | | |
| Cluster 3 vs 1* | 0.44 (0.23-0.84) | 0.013 | | |
| Age (years) | 1.01 (0.98-1.04) | 0.53 | | |
| Female | 1.34 (0.84-2.15) | 0.22 | | |
| BMI (kg/m$^2$) | 0.98 (0.94-1.01) | 0.18 | | |
| NYHA classification 3-4 | 1.78 (1.12-2.82) | 0.015 | 1.70 (1.05-2.77) | 0.033 |
| Atrial fibrillation | 1.28 (0.78-2.09) | 0.33 | | |
| Diabetes | 1.49 (0.95-2.36) | 0.08 | 1.45 (0.89-2.36) | 0.13 |
| Ischemic etiology | 0.91 (0.56-1.45) | 0.68 | | |
| COPD | 2.21 (1.15-4.25) | 0.017 | 2.42 (1.21-4.86) | 0.012 |
| LA volume indexed (ml/m$^2$) | 1.01 (0.99-1.02) | 0.38 | | |
| E/e’ septal ratio | 1.03 (0.99-1.06) | 0.064 | 1.02 (0.99-1.05) | 0.21 |
| eSPAP (mmHg) | 1.02 (0.99-1.04) | 0.14 | | |
| Hemoglobin (g/dL) | 0.89 (0.79-1.01) | 0.08 | 0.97 (0.85-1.12) | 0.68 |
| eGFR (mL/min/1.73m$^2$) by CK-EPI | 0.98 (0.97-0.99) | 0.002 | 0.98 (0.97-0.99) | 0.020 |
| NT-proBNP (log pg/mL) | 1.18 (0.95-1.47) | 0.13 | | |

BMI, body mass index; COPD, chronic obstructive pulmonary disease; eGFR, estimated glomerular filtration rate; eSPAP, estimated systolic pulmonary artery pressure; HF, heart failure; LA, left atrium; NT-proBNP, N-terminal pro B-type natriuretic peptide; NYHA, New York Heart Association

*Not included in the multivariable model

**Supplementary Table S5.** Targeted analysis of plasma amino and organic acids in a subset of non-HF and HFpEF subjects from the secondary Canadian cohort.

| | Non-HF | HFpEF | Fold-change | *P value* |
|---|---|---|---|---|
| | N= 103 | N=74 | | |
| **Amino acids (µM)** | | | | |
| Alanine | 504.7 ± 106.2 | 527.5 ± 126.8 | 1.02 | 0.502 |
| Arginine | 163.7 ± 38.2 | 161.0 ± 51.9 | 0.98 | 0.689 |
| Asparagine | 47.8 ± 10.5 | 44.8 ± 10.4 | 0.94 | 0.157 |
| Aspartate | 5.2 ± 1.5 | 5.4 ± 1.5 | 1.03 | 0.516 |
| Cysteine | 324.2 ± 47.3 | 369.1 ± 74.4 | 1.10 | $4.93 \times 10^{-5}$ |
| Glutamate | 76.8 ± 33.3 | 86.0 ± 39.9 | 1.09 | 0.160 |
| Glutamine | 598.7 ± 75.0 | 576.7 ± 77.1 | 0.96 | 0.050 |
| Glycine | 262.7 ± 60.3 | 245.9 ± 48.1 | 0.95 | 0.144 |
| Histidine | 95.4 ± 25.3 | 90.3 ± 28.8 | 0.93 | 0.167 |
| **Hydroxyproline** | **2.7 ± 0.9** | **3.4 ± 2.1** | **1.19** | **0.003** |
| Isoleucine | 64.3 ± 19.6 | 67.1 ± 22.2 | 1.04 | 0.381 |
| Leucine | 138.4 ± 39.4 | 140.8 ± 44.9 | 1.01 | 0.881 |
| Lysine | 199.0 ± 38.4 | 202.7 ± 37.1 | 1.03 | 0.303 |
| Methionine | 27.2 ± 6.7 | 27.1 ± 7.4 | 0.99 | 0.732 |
| Phenylalanine | 62.8 ± 12.3 | 66.7 ± 14.3 | 1.05 | 0.100 |
| Proline | 257.0 ± 84.2 | 266.2 ± 73.6 | 1.03 | 0.471 |
| Serine | 156.2 ± 36.4 | 142.6 ± 37.5 | 0.91 | 0.013 |
| Threonine | 140.0 ± 36.9 | 132.7 ± 33.2 | 0.96 | 0.330 |
| Tryptophan | 55.2 ± 27.4 | 49.3 ± 13.7 | 0.92 | 0.038 |
| Tyrosine | 86.3 ± 22.6 | 86.0 ± 27.0 | 0.99 | 0.860 |
| Valine | 254.3 ± 56.5 | 258.4 ± 62.3 | 1.01 | 0.741 |
| **Organic acids (µM)** | | | | |
| β-Hydroxyisobutyrate | 18.4 ± 9.3 | 21.5 ± 10.1 | 1.16 | 0.036 |
| Acetoacetate | 27.8 ± 26.6 | 28.0 ± 13.7 | 1.07 | 0.358 |
| α-Hydroxybutyrate | 687.8 ± 304.4 | 683.8 ± 263.6 | 1.02 | 0.787 |
| α-Hydroxyglutarate | 8.3 ± 2.3 | 9.2 ± 6.0 | 1.03 | 0.477 |
| α-Ketobutyrate | 6.2 ± 3.4 | 5.7 ± 3.2 | 0.93 | 0.419 |
| β-Hydroxybutyrate | 46.3 ± 77.1 | 52.0 ± 45.1 | 1.19 | 0.094 |
| Citrate | 105.0 ± 20.6 | 115.3 ± 26.3 | 1.06 | 0.064 |
| **α-Ketoglutarate** | **54.1 ± 13.5** | **65.8 ± 20.2** | **1.18** | **$2.52 \times 10^{-5}$** |
| Lactate | 1399.1 ± 844.7 | 1617.8 ± 787.5 | 1.15 | 0.050 |
| **Malate** | **7.6 ± 2.1** | **9.7 ± 3.8** | **1.20** | **$5.18 \times 10^{-5}$** |
| Pyruvate | 707.6 ± 268.6 | 738.6 ± 242.3 | 1.03 | 0.533 |
| Succinate | 21.7 ± 14.0 | 21.2 ± 7.9 | 1.03 | 0.476 |
| **Calculated ratios** | | | | |
| Lactate/Pyruvate | 2.3 ± 2.3 | 2.5 ± 2.2 | 1.11 | 0.166 |
| β-Hydroxybutyrate/Acetoacetate | 1.5 ± 0.7 | 1.8 ± 0.9 | 1.12 | 0.108 |
| α-Hydroxybutyrate/α-Ketobutyrate | 151.5 ± 157.3 | 157.1 ± 127.3 | 1.10 | 0.233 |
| Glutamine/Glutamate | 9.1 ± 3.6 | 8.1 ± 3.7 | 0.88 | 0.057 |

Values are mean ± standard deviation. Metabolites shown in this table had ≤ 20% missing values within each group (i.e. HFpEF or controls). Missing values were imputed as 90% of the minimal value of each metabolite. Fold-changes and *P*-values were calculated using linear regression with batch, age and sex as covariates. Metabolite shown in bold are statistically significant for HFpEF vs. Non-HF using $P \leq 0.01$ and absolute fold-change │FC │>1.15 as threshold.

**Supplementary Table S6. Baseline characteristics in HFpEF patients of the Canadian secondary cohort according to cluster subgroup.**

| | Cluster 1 | Cluster 2 | Cluster 3 | *P (Levene)* | *P (Anova)* | *P (Kruskal-Wallis)* |
|---|---|---|---|---|---|---|
| | N=18 | N=25 | N=31 | | | |
| Age (years) | 70.9 ± 6.6 | 69.8 ± 6.0 | 70.2 ± 7.8 | 0.408 | 0.875 | 0.926 |
| Female (*n*, %) | 6 (33) | 10 (40) | 12 (39) | N/A | N/A | 0.898 (chi-square) |
| BMI | 33.6 ± 5.6 | 33.5 ± 6.6 | 31.3 ± 5.6 | 0.760 | 0.287 | 0.397 |
| Cholesterol (mmol/L) | 3.8 ± 0.6 | 4.0 ± 0.8 | 3.9 ± 0.9 | 0.611 | 0.633 | 0.516 |
| **LDL (mmol/L)** | **1.6 ± 0.4[#†]** | **2.1 ± 0.7** | **2.0 ± 0.6** | **0.376** | **0.023** | **0.026** |
| **HDL (mmol/L)** | **0.9 ± 0.2[†]** | **0.9 ± 0.3[†]** | **1.2 ± 0.4** | **0.378** | **$2.63x10^{-6}$** | **$3.36x10^{-6}$** |
| **Triglycerides (mmol/L)** | **3.5 ± 1.3[#†]** | **2.5 ± 0.6[†]** | **1.3 ± 0.3** | **0.084** | **$4.19x10^{-18}$** | **$3.09x10^{-12}$** |
| Glucose (mmol/L) | 7.6 ± 2.6 | 8.9 ± 3.8[†] | 7.1 ± 2.5 | 0.811 | 0.084 | 0.101 |
| Creatinine (umol/L) | 109.2 ± 34.6[†] | 117.5 ± 54.5 | 92.2 ± 31.3 | 0.082 | 0.069 | 0.054 |
| eGFR (ml/min/1.73m$^2$) | 59.3 ± 19.3 | 59.0 ± 24.1 | 70.1 ± 20.7 | 0.170 | 0.107 | 0.159 |

Values are mean ± standard deviation. Statistic tests were performed on log2-transformed data. Equality of variance between clusters was tested by Levene test. Variables shown in bold are statistically significant with P≤0.05 following Kruskal-Wallis test (or chi-squared test where appropriate).

\# p<0.05 individual category vs. cluster 2 following Wilcoxon test

† p<0.05 individual category vs. cluster 3 following Wilcoxon test

**Supplementary Table S7.** Targeted analysis of plasma amino and organic acids in HFpEF patients of the Canadian cohort according to cluster subgroups.

| | Cluster 1 | Cluster 2 | Cluster 3 | *P (Levene)* | *P (Anova)* | *P (Kruskal-Wallis)* |
|---|---|---|---|---|---|---|
| | N=18 | N=25 | N=31 | | | |
| **Amino acids (µM)** | | | | | | |
| Alanine | 357.8 ± 87.4 | 377.3 ± 108.1† | 360.4 ± 125.0 | 0.990 | 0.054 | 0.055 |
| Arginine | 187.5 ± 71.2 | 203.2 ± 87.0 | 200.1 ± 54.9 | 0.516 | 0.201 | 0.329 |
| Asparagine | 38.8 ± 8.2 | 39.2 ± 9.7 | 36.5 ± 11.2 | 0.092 | 0.544 | 0.321 |
| Aspartate | 5.8 ± 3.6† | 4.8 ± 2.1 | 5.0 ± 2.7 | 0.421 | 0.131 | 0.118 |
| **Cysteine** | **217.8 ± 80.7†** | **213.2 ± 64.2†** | **232.6 ± 95.9** | **0.387** | **0.008** | **0.006** |
| **Glutamate** | **122.4 ± 113.0#†** | **93.7 ± 51.6** | **114.9 ± 106.8** | **0.590** | **0.001** | **0.001** |
| Glutamine | 518.5 ± 122.8 | 558.4 ± 105.3 | 501.4 ± 141.1 | 0.837 | 0.363 | 0.401 |
| Glycine | 238.4 ± 72.4 | 220.1 ± 57.4 | 226.5 ± 61.6 | 0.518 | 0.315 | 0.201 |
| Histidine | 70.2 ± 9.1 | 73.0 ± 11.9 | 67.1 ± 12.4 | 0.079 | 0.401 | 0.528 |
| Hydroxyproline | 3.0 ± 1.9 | 2.6 ± 0.8 | 2.7 ± 1.1 | 0.556 | 0.920 | 0.771 |
| Isoleucine | 79.8 ± 23.4† | 80.5 ± 29.4 | 91.2 ± 42.3 | 0.907 | 0.112 | 0.092 |
| Leucine | 111.3 ± 32.6† | 110.2 ± 36.0 | 120.2 ± 46.9 | 0.782 | 0.220 | 0.096 |
| Lysine | 201.3 ± 56.8 | 202.3 ± 57.7 | 198.9 ± 62.9 | 0.362 | 0.688 | 0.694 |
| Methionine | 26.0 ± 9.4 | 23.3 ± 6.8 | 25.5 ± 10.2 | 0.221 | 0.546 | 0.805 |
| Phenylalanine | 69.1 ± 15.6 | 67.9 ± 12.4 | 68.4 ± 15.5 | 0.612 | 0.520 | 0.258 |
| **Proline** | **232.8 ± 66.9†** | **235.9 ± 69.7†** | **241.1 ± 91.9** | **0.450** | **0.003** | **0.002** |
| Serine | 93.0 ± 26.8† | 83.6 ± 16.4 | 79.7 ± 20.9 | 0.012 | 0.205 | 0.159 |
| Threonine | 135.3 ± 44.5 | 125.7 ± 41.9 | 117.5 ± 37.4 | 0.428 | 0.902 | 0.876 |
| Tryptophan | 55.8 ± 22.0 | 55.7 ± 19.6 | 57.3 ± 21.6 | 0.981 | 0.201 | 0.345 |
| Tyrosine | 76.6 ± 27.9 | 73.4 ± 23.5 | 72.2 ± 25.1 | 0.671 | 0.372 | 0.357 |
| **Valine** | **242.1 ± 67.2†** | **243.3 ± 61.6** | **266.5 ± 71.2** | **0.775** | **0.074** | **0.021** |
| **Organic acids (µM)** | | | | | | |
| β-Hydroxyisobutyrate | 25.4 ± 11.4† | 21.9 ± 10.6 | 18.8 ± 8.3 | 0.376 | 0.148 | 0.088 |
| Acetoacetate | 7.9 ± 8.7 | 11.9 ± 18.1 | 15.4 ± 25.8 | 0.540 | 0.689 | 0.477 |
| α-Hydroxybutyrate | 1351.9 ± 675.9 | 1351.9 ± 614.1 | 1644.2 ± 891.7 | 0.090 | 0.813 | 0.544 |
| α-Hydroxyglutarate | 9.3 ± 4.2 | 9.3 ± 3.4 | 9.1 ± 8.3 | 0.678 | 0.450 | 0.109 |
| α-Ketobutyrate | 3.9 ± 3.3 | 3.9 ± 1.4 | 5.8 ± 3.6 | 0.942 | 0.724 | 0.666 |
| β-Hydroxybutyrate | 79.1 ± 104.7 | 94.1 ± 110.2 | 82.7 ± 87.7 | 0.238 | 0.477 | 0.414 |
| Citrate | 98.1 ± 27.8 | 107.7 ± 28.5 | 107.7 ± 35.2 | 0.448 | 0.728 | 0.609 |
| α-Ketoglutarate | 30.8 ± 12.4 | 34.4 ± 13.8 | 36.1 ± 15.4 | 0.650 | 0.823 | 0.743 |
| Lactate | 1944.1 ± 920.5 | 1830.9 ± 880.1 | 1644.2 ± 794.3 | 0.465 | 0.414 | 0.385 |
| **Malate** | **10.3 ± 2.2†** | **9.9 ± 3.5†** | **8.9 ± 2.0** | **0.743** | **0.057** | **0.031** |
| Pyruvate | 241.4 ± 126.3 | 296.2 ± 144.2 | 273.8 ± 130.2 | 0.300 | 0.164 | 0.217 |
| Succinate | 19.4 ± 5.6 | 18.3 ± 3.0† | 17.7 ± 3.0 | 0.029 | 0.042 | 0.074 |
| **Calculated ratios** | | | | | | |
| Lactate/Pyruvate | 16.5 ± 20.7 | 7.3 ± 4.6 | 7.0 ± 4.2 | 0.657 | 0.928 | 0.185 |
| β-Hydroxybutyrate/Acetoacetate | 46.6 ± 88.9 | 54.3 ± 102.1 | 59.6 ± 102.6 | 0.627 | 0.548 | 0.620 |
| α-Hydroxybutyrate/α-Ketobutyrate | 745.0 ± 864.2 | 369.6 ± 172.1 | 323.1 ± 125.3 | 0.088 | 0.884 | 0.416 |
| **Glutamine/Glutamate** | **7.8 ± 5.6#†** | **8.0 ± 4.7** | **8.5 ± 6.2** | **0.713** | $2.51 \times 10^{-4}$ | **0.001** |

Values are mean ± standard deviation. Missing values were imputed as 90% of the minimal value of each metabolite. Statistic tests were performed on log2-transformed data. Metabolites shown in bold are statistically significant with P≤0.05 following Kruskal-Wallis test.

\# p<0.05 individual category vs. cluster 2 following Wilcoxon test
† p<0.05 individual category vs. cluster 3 following Wilcoxon test
